# PAUSE-Agents: A Clinician-in-the-Loop Multi-Agent AI Pipeline for ICU-to-Ward Handoff Briefs

**DOI:** 10.64898/2026.07.10.26357759

**Authors:** Saki Amagai, Wan-Ting Liao, Caleb Murphy, Courtney Reamer, Yupeng Liu, Bhavana Ambil, Glenn Fernandes, Lekshmi Santhosh, Patrick G Lyons, Neil Jordan, David Liebovitz, Adrienne Kline, Juan C Rojas, Yuan Luo, Catherine A. Gao

## Abstract

ICU-to-ward transfers are high-risk transitions marked by information loss and burdensome handoff preparation. We developed PAUSE-Agents, a clinician-in-the-loop multi-agent LLM pipeline that drafts source-attributed handoff briefs from structured ICU data and clinical notes using the clinician-developed ICU-PAUSE template. Mirroring ICU team structure, PAUSE-Agents routes each record through a scribe extractor, 6 role-specialized agents, explicit conflict surfacing, and deterministic safety checks before synthesis, producing an editable first draft rather than an autonomous note. In a single-center medical ICU cohort, 5 physicians completed 100 reviews of 84 agent-drafted briefs. Among adjudicable claims, 98.8% were verified and 1.2% were incorrect; 88% of briefs had no pertinent omission, and mean PDSQI-9 quality was 4.20/5. PAUSE-Agents surfaced 118 conflict warnings and 421 safety flags, making documentation inconsistencies visible before handoff. An o4-mini PDSQI-9 judge showed limited case-level discrimination but supported aggregate monitoring. We release PAUSE-Agents and its clinician evaluation application.

## Introduction

Hospital intensive care unit (ICU)-to-ward transfers are not a single handoff, but a complex transition process.^1^ At transition points, responsibility shifts across teams through overlapping exchanges among physicians, nurses, respiratory therapists, pharmacists, case managers, rehabilitation therapists, and others. The accepting team must quickly grasp why the patient required critical care, what changed in the ICU, which diagnostic questions remain open, which medications or devices require active follow-up, and what goals-of-care constraints should guide escalation. When this information is incomplete, buried in the chart, or internally inconsistent, preventable harm can follow.^2,3^ While general handoff frameworks such as I-PASS demonstrate that structured communication can improve handoff quality and safety,^4^ ICU-to-ward-specific tools such as ICU-PAUSE define the information elements needed for this particular transition.^5–9^ However, even well-designed handoff tools cannot fully eliminate information loss across heterogeneous handoff workflows. Simpler informatics approaches, such as electronic health record (EHR) templates and automated data-filling, may improve capture of predefined fields but are limited in determining clinical salience, reconciling internal inconsistencies, explaining why a finding appears, or formulating a transfer plan; they may also contribute to note bloat (extraneous, irrelevant, or duplicated information) and copy-forward risk.^10,11^

Large language models (LLMs) are an appealing candidate for this problem: they can read multi-document EHR data and draft fluent clinical summaries at scale, and recent work in clinical summarization and discharge documentation shows that LLM-generated summaries can be useful and, in selected settings, comparable with physician-authored summaries.^12–17^ Yet ICU-to-ward handoff is a high-stakes summarization task in which omissions, unsupported assertions, and poorly reconciled medication or code-status information can directly affect care. Most current LLM summarization workflows still rely on a single model pass over the full clinical record —multiple clinical note types together with structured EHR data —a design vulnerable to hallucination, lost-in-the-middle effects, and the conflation of distinct clinical reasoning tasks such as ventilator-status assessment, medication reconciliation, diagnostic uncertainty, and disposition planning.^18^

Agentic artificial intelligence (AI) systems address some of these limitations by decomposing complex work into bounded subtasks, connecting LLMs to external tools, and allowing intermediate outputs to be checked before final synthesis. Early medical-agent systems have shown promise in diagnosis, longitudinal management, triage, and clinical decision support.^19–21^ Recent benchmarking, however, cautions that generic agentic designs can add substantial token and latency cost while yielding only modest accuracy gains and hallucination.^22,23^ The relevant question for clinical deployment is, therefore, not whether agents are categorically better than single LLMs, but whether a task-grounded decomposition creates safety-relevant capabilities that a monolithic prompt does not naturally provide.

ICU-to-ward transfer documentation is well matched to such a decomposition because the predictable failure modes of ICU-to-ward transfers (e.g., medication reconciliation, respiratory trajectory, code status, disposition, and pending diagnostics) map onto distinct clinical perspectives.^24,25^ The intensivist integrates the global illness course, while the bedside nurse, respiratory therapist, pharmacist, dietitian, case manager, and rehabilitation therapist each observe a different facet of the same patient. Existing agentic summarization systems generally draft or refine a single longitudinal narrative; the most similar published hospital-course summarization workflow uses sequential draft-and-review steps rather than parallel, role-specialized clinical perspectives.^14^ To our knowledge, no prior system has applied a team-mirroring multi-agent architecture to ICU summarization or the ICU-to-ward transfer note—the artifact whose purpose is precisely to reconcile these perspectives into a single handoff. The value here is not better fluency, but independent role-scoped extraction followed by explicit conflict surfacing before synthesis.

Here we present PAUSE-Agents, a source-attributed, clinician-in-the-loop, multi-agent pipeline that generates ICU-to-ward handoff briefs from structured data in the Common Longitudinal ICU Format (CLIF)^26^ and routed clinical notes as illustrated in **Figure 1**. The briefs are formatted according to the clinician-developed ICU-PAUSE handoff template, a structured mnemonic spanning ICU course, code status, areas of uncertainty, pending tests, active consultants, high-risk-medication unprescribing, a problem/to-do summary, and the transfer exam. PAUSE-Agents operationalizes the team-mirroring design through independent role-scoped views, deterministic safety checks, clinician-facing conflict-warning banners, and clinician-in-the-loop editing of a rendered first draft rather than autonomous note generation. We evaluated three questions: whether the generated briefs met a physician-assessed clinical bar for accuracy, completeness, and usefulness; whether independent specialist views surfaced cross-domain documentation conflicts for clinician review before finalizing the handoff; and whether an automated LLM judge using the nine-item Provider Documentation Summarization Quality Instrument (PDSQI-9) could provide a scalable, partially validated monitor of brief quality.^27,28^

**Figure 1.**
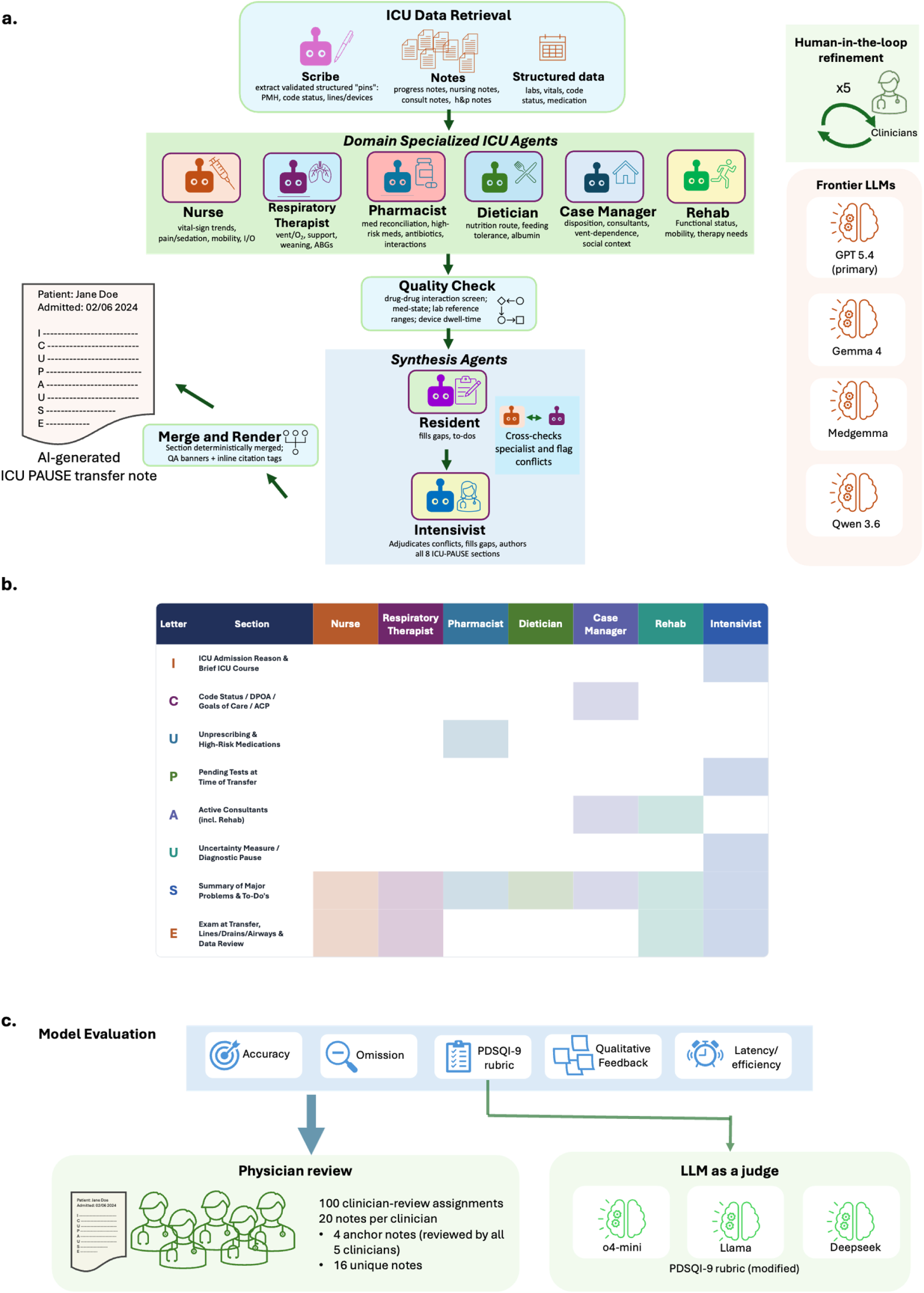
PAUSE-Agents generation pipeline, agent-section ownership, and evaluation framework. (a) Generation pipeline. PAUSE-Agents retrieves structured CLIF data and routes clinical notes anchored to the ICU-to-ward transfer, then passes role-scoped inputs through a structured-extraction scribe and 6 role-specialized clinical agents: nurse, respiratory therapist, pharmacist, dietitian, case manager, and rehabilitation. A deterministic safety layer checks rule-verifiable issues before resident and intensivist synthesis. The final ICU-PAUSE brief is deterministically merged and rendered with inline source-citation tags and clinician-facing warning banners as an editable first draft. (b) Agent-section ownership. Rows show the 8 ICU-PAUSE handoff sections and columns show contributing agents; shaded cells indicate primary ownership or contribution, with the intensivist harmonizing all sections at synthesis. (c) Evaluation framework. The pipeline was refined in a 25-case, human-in-the-loop development phase and then evaluated on 84 held-out briefs. Five physicians completed 100 structured reviews, with 4 anchor briefs reviewed by all 5 physicians for inter-rater reliability. Primary outcomes were sentence-level claim accuracy and per-domain pertinent omission; PDSQI-9 quality was secondary. An o4-mini PDSQI-9 judge was evaluated as an exploratory monitor against physician-counted errors and omissions.

### Results Study corpus

The study corpus comprised a development/human-in-the-loop (HITL) set of 25 ICU-to-ward transfer notes used for iterative pipeline refinement and a held-out summative set of 84 transfer notes used for final evaluation (**Table 1**). The unit of analysis was the ICU-to-ward handoff brief. Because a single hospitalization may involve more than one ICU-to-ward transfer, we included only the first transfer per hospitalization. Therefore, each hospitalization contributed one transfer note, and no duplicate transfer notes from the same hospitalization entered the analysis. Eligible source notes comprised 8 types — history and physical (H&P), progress, consult, plan-of-care, nursing, case management, social work, and therapy notes – routed to agents. These are subsequently routed to agents by role, with the admission H&P routed to all (routing detail in **Methods**).

**Table 1.**
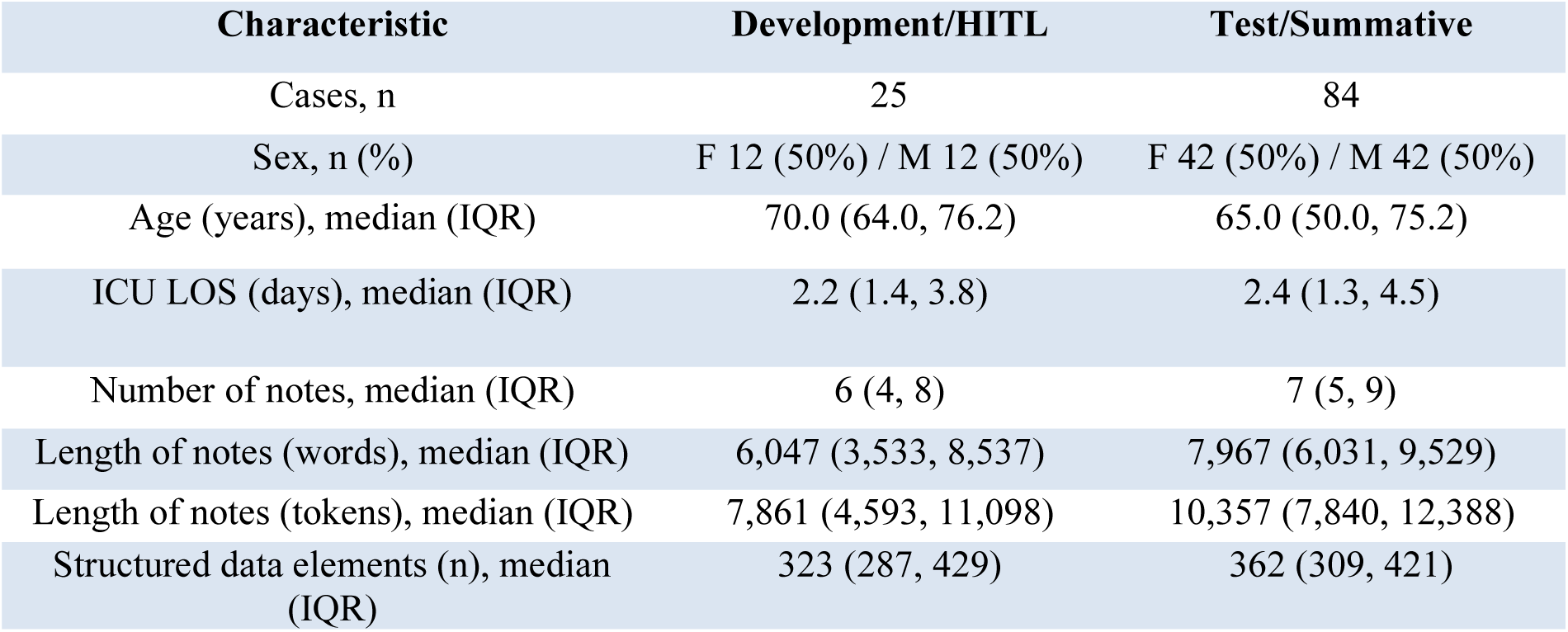
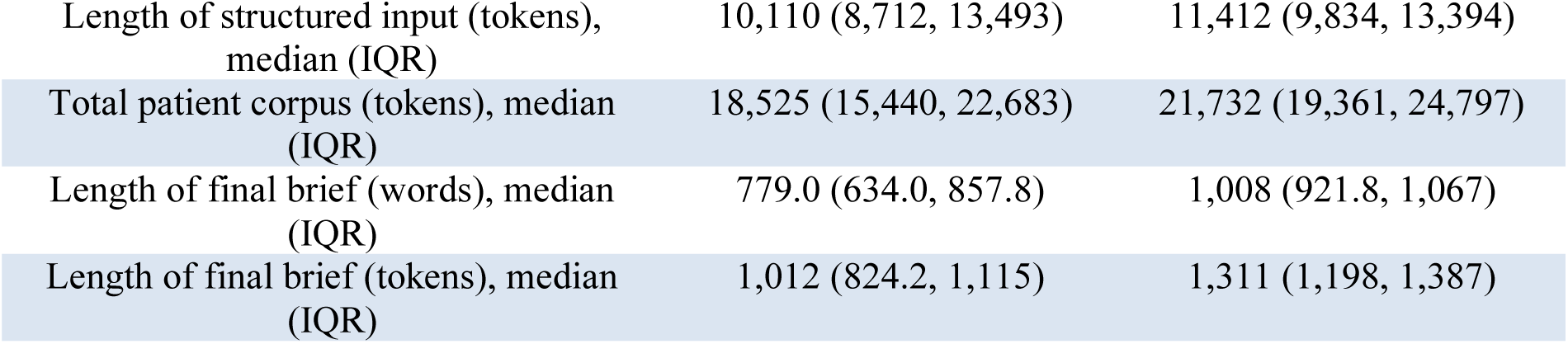
Study corpus characteristics. The development/Human-in-the-Loop (HITL) set comprised 25 ICU-to-ward transfer notes reviewed across 5 iterative pilot rounds, and the held-out test/summative set comprised 84 ICU-to-ward transfer notes used for final evaluation and not seen during development; descriptive statistics are reported over the de-duplicated brief set. All cases were from the MICU; provider specialty was not stratified. Token lengths were estimated with the tiktoken cl100k_base encoder for consistent corpus-size comparison across cohorts, whereas per-agent counts in Supplementary Figure S1 reflect the production GPT-5.4 tokenizer recorded at run time. P-values were calculated using the Mann–Whitney U test for continuous variables and Fisher’s exact test for sex.

| Characteristic | Development/HITL | Test/Summative |
| --- | --- | --- |
| Cases, n | 25 | 84 |
| Sex, n (%) | F 12 (50%) / M 12 (50%) | F 42 (50%) / M 42 (50%) |
| Age (years), median (IQR) | 70.0 (64.0, 76.2) | 65.0 (50.0, 75.2) |
| ICU LOS (days), median (IQR) | 2.2 (1.4, 3.8) | 2.4 (1.3, 4.5) |
| Number of notes, median (IQR) | 6 (4, 8) | 7 (5, 9) |
| Length of notes (words), median (IQR) | 6,047 (3,533, 8,537) | 7,967 (6,031, 9,529) |
| Length of notes (tokens), median (IQR) | 7,861 (4,593, 11,098) | 10,357 (7,840, 12,388) |
| Structured data elements (n), median (IQR) | 323 (287, 429) | 362 (309, 421) |
| Length of structured input (tokens), median (IQR) | 10,110 (8,712, 13,493) | 11,412 (9,834, 13,394) |
| Total patient corpus (tokens), median (IQR) | 18,525 (15,440, 22,683) | 21,732 (19,361, 24,797) |
| Length of final brief (words), median (IQR) | 779.0 (634.0, 857.8) | 1,008 (921.8, 1,067) |
| Length of final brief (tokens), median (IQR) | 1,012 (824.2, 1,115) | 1,311 (1,198, 1,387) |

Both sets were drawn from the same single-center medical ICU (MICU) population and were comparable in sex, age, and ICU length of stay (all P > 0.2; **Table 1**). The summative set was larger and contained longer notes than the development set, so held-out performance was evaluated under a more demanding long-context condition rather than an easier matched sample. Each summative record was a multi-document summarization task, with a median of 7 clinical notes, 10,357 note tokens, and 362 structured data elements per patient, corresponding to a median total input size of 21,732 tokens. From these inputs, PAUSE-Agents produced ICU-PAUSE briefs with a median length of 1,008 words, or 1,311 tokens, corresponding to an approximately 17-fold compression of the source record into the standardized handoff format.

### Evaluation framework

For each patient, routed clinical notes and structured CLIF data were used to generate a single ICU-to-ward handoff brief in the clinician-developed ICU-PAUSE format.^26^ We evaluated each brief along three dimensions: factual accuracy, defined as the proportion of adjudicable claims supported by the source record; completeness, defined as the absence of pertinent omissions across nine clinical data domains; and perceived documentation quality, measured with PDSQI-9.^28^ Accuracy and completeness were the primary outcomes, and PDSQI-9 quality was the secondary outcome.

### Physician evaluation of brief quality

Five physician reviewers (2 hospitalist attendings, 2 internal medicine residents, and 1 pulmonary critical care attending) completed 100 structured reviews of the 84-note summative cohort. Eighty briefs were reviewed by 1 clinician, and 4 anchor briefs were reviewed by all 5 clinicians for inter-rater reliability, yielding 16 unique briefs plus 4 shared anchors per reviewer. Structured review required a median of 10.5 minutes per case and was the rate-limiting step of the evaluation (see Evaluation efficiency). The assignment design was informed by the HITL phase, in which reviewers showed high concordance on objective accuracy and omission judgments, and balanced targeted inter-rater overlap with broader held-out case coverage. Near-perfect agreement on the summative anchor cases supported pooling single-review claim- and omission-level judgments across the full cohort. For claim- and omission-level analyses, all 100 physician reviews were included. For brief-level PDSQI-9 analyses, the 5 ratings for each anchor brief were averaged into a single per-brief value so that each of the 84 held-out briefs contributed once.

#### Accuracy (primary outcome)

Physicians evaluated each brief at the sentence level, marking each atomic, source-checkable factual claim as verified, incorrect, or cannot verify against the source record. Across 5,918 reviewed claims, 5,754 were verified, 70 were judged incorrect, and 94 were marked cannot verify. Cannot-verify labels largely reflected reviewer-facing de-identification of identity-dependent fields rather than deployment conditions (**Methods**). Excluding cannot-verify claims from the adjudicable denominator, 98.8% of claims were supported by the source record (5,754/5,824), and 1.2% were incorrect (70/5,824); 62% of briefs contained no incorrect claim (52/84). Clinicians showed near-perfect agreement on these verdicts across anchor cases (Gwet’s AC2, 0.98–0.99; see Inter-rater reliability). Errors were concentrated in the most synthesis-dependent sections: ICU Admission/Course (95.7% accurate) and Unprescribing/High-Risk Medications (97.0%) carried the highest error rates, whereas the remaining sections each exceeded 98.5% accuracy (**Figure 3a; Supplementary Table S2**). This pattern suggests that future gains may come from improving temporal event ordering and medication-state reconciliation rather than generic summarization fluency.

**Figure 2.**
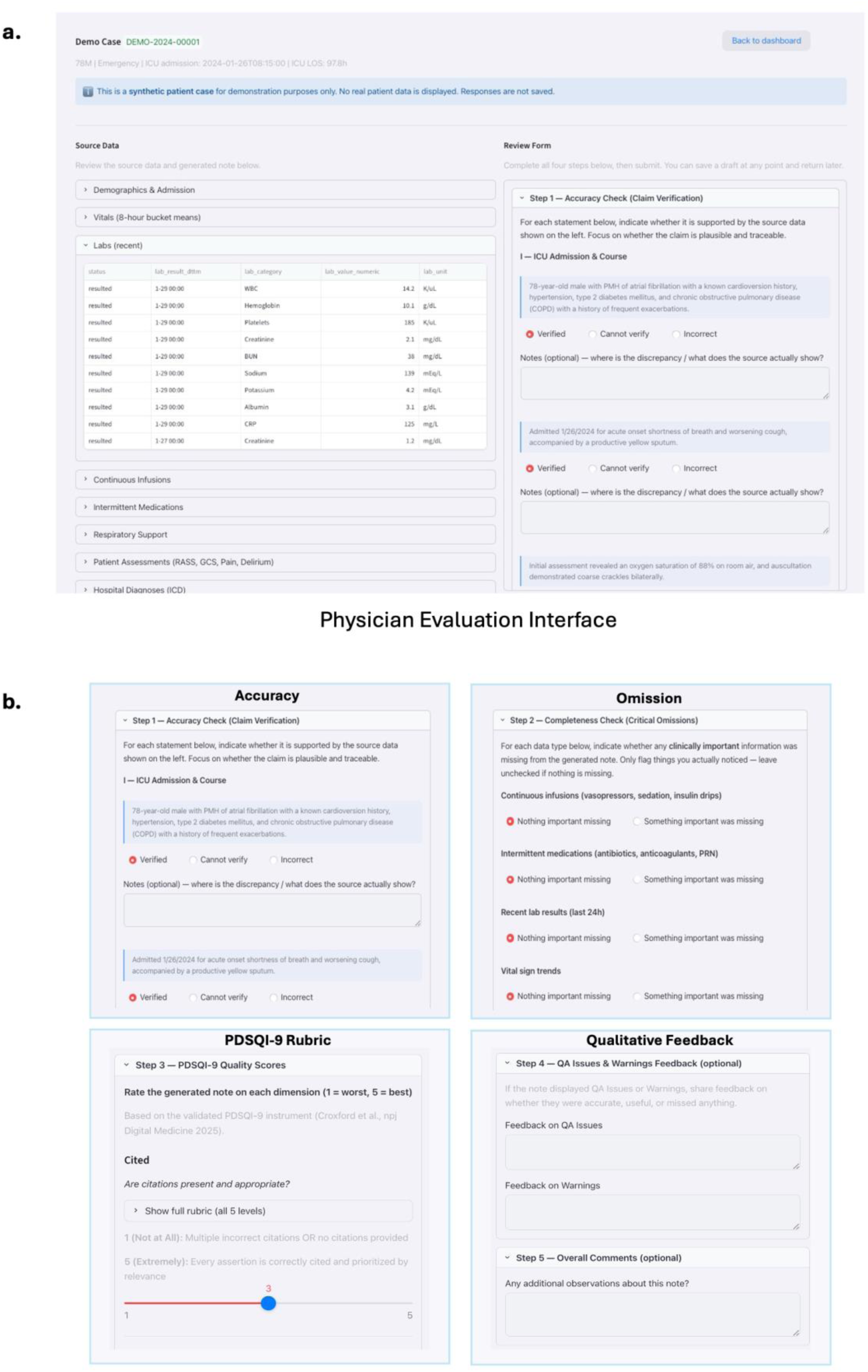
PHI-compliant clinician review application. Screenshots of the web interface used for human evaluation, shown with a synthetic demo case and no protected health information (PHI). (a) Overall review layout: the generated PAUSE-Agents brief is displayed alongside the structured source data and routed clinical notes (left), with the structured review form (right); the rendered ICU-Pause briefs that reviewers evaluated was displayed in the lower left of the same interface and is not shown in the screenshot (b) Close-up of the four review components completed for each brief: Accuracy (Step 1, sentence-level claim verification and adjudication), Omission (Step 2, per-domain completeness assessment), PDSQI-9 Rubric (Step 3, documentation-quality scoring using the validated PDSQI-9 anchors), and Qualitative Feedback (Step 4, feedback on surfaced QA warnings, and Step 5, overall free-text comments). The application ran within the institution’s HIPAA-compliant environment behind Microsoft Entra ID single sign-on. The same generated brief and source bundle were scored in parallel by the automated o4-mini PDSQI-9 judge, whose validation against physician judgments is shown in Figure 4.

**Figure 3.**
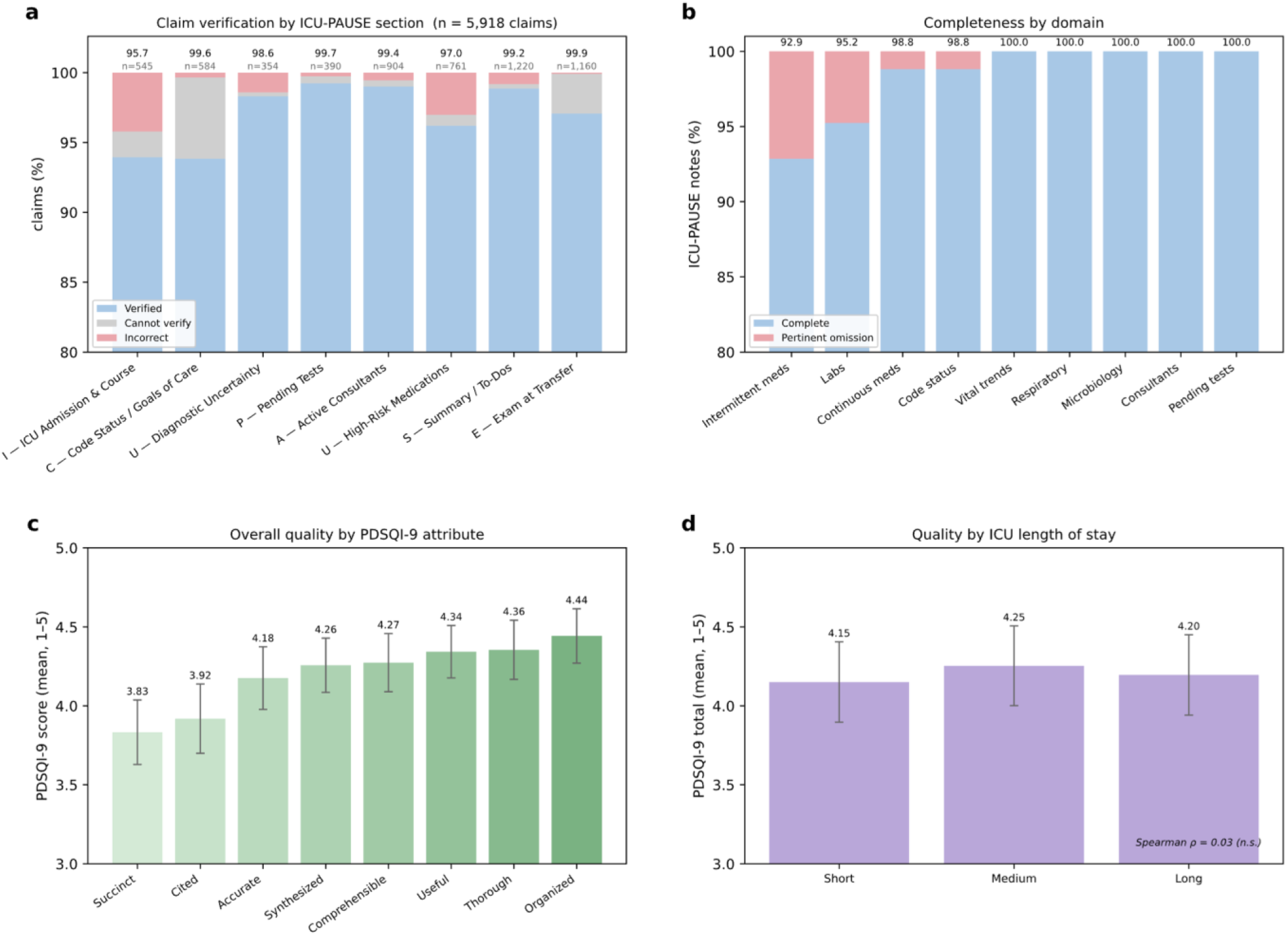
Physician evaluation of brief quality. Physician evaluation of deployed GPT-5.4 PAUSE-Agents briefs across the 84-case summative cohort. Panels a and b pool all 100 physician reviews; panels c and d are computed at the brief level, with multiply reviewed briefs averaged into a single per-brief value. (a) Sentence-level claim verification by ICU-PAUSE section, showing verified, cannot verify, and incorrect claims. Numerals indicate section accuracy and claim count. (b) Completeness across 9 clinical data domains, showing briefs with no pertinent omission versus those with an omission. (c) Mean physician PDSQI-9 scores across the 8 Likert attributes; the binary stigmatizing-language item, flagged in 0% of briefs, is omitted. (d) Mean PDSQI-9 total score stratified by ICU length of stay, showing no quality degradation with longer stays. In a and b, note the y-axis is truncated to the 80-100% range for visibility to show small non-verified and omitted fractions; in c and d, error bars indicate 95% confidence intervals.

#### Completeness (primary outcome)

Reviewers assessed whether each brief omitted important information across 9 clinical data domains. Pertinent omissions were uncommon: 88% of briefs had no omission flagged, and domain-level completeness was ≥92.9% across all domains (**Figure 3b; Supplementary Table S3**). Inter-clinician agreement on omission judgments was near-perfect on anchor cases (Gwet’s AC2 = 0.97). Omissions were concentrated in intermittent medications (92.9% complete; 6 omissions) and recent laboratory data (95.2% complete; 4 omissions), while the remaining domains had no or near-zero omissions. The intermittent-medication shortfall largely reflected limited chronic oral-medication coverage in the source data rather than model misses, whereas laboratory omissions were rare.

#### Overall quality — PDSQI-9 (secondary outcome)

Physician PDSQI-9 ratings averaged 4.20/5 across the 8 Likert attributes **(Figure 3c; Supplementary Table S1**), and no brief was flagged for stigmatizing language. Scores were highest for organization, thoroughness, and usefulness, and lowest for succinctness and citation reviewability. The lower citation-reviewability score likely reflected the de-identification workflow rather than citation absence alone: Philter occasionally over-redacted the source notes displayed to reviewers, obscuring the text to which some citation tags pointed and making those citations harder to trace. Cited was also among the least reliably rated attributes both in the original PDSQI-9 validation and in our anchor set (AC2 0.61).

#### Inter-rater reliability

Agreement among the 5 physicians was near-perfect for the primary objective measures. Across the four anchor briefs, Gwet’s AC2 was 0.98 for three-way claim verdicts, 0.99 for binary correct-versus-incorrect judgments, and 0.97 for omission judgments (**Supplementary Table S4**). Reviewer identity explained little variance in incorrect-claim rate or pertinent-omission count, supporting the use of pooled single-review judgments for the primary outcomes. Agreement on subjective PDSQI-9 ratings was lower and more sensitive to reviewer leniency, supporting our treatment of claim-level accuracy and omissions as primary outcomes and PDSQI-9 as secondary.

#### Quality versus length of stay

Brief quality did not degrade with ICU length of stay on any evaluated axis, consistent with the fixed 48-hour lookback window.

#### Qualitative clinician feedback

Optional free-text comments, provided by 3 of 5 reviewers, were generally favorable and identified both strengths and areas for refinement (**Supplementary Table S5**). Reviewers praised the briefs’ thoroughness, organization, transfer planning, and synthesis of complex ICU courses, including anticipatory guidance, medication-stewardship recommendations, and explicit diagnostic uncertainty. Suggested refinements focused mainly on prioritization and completeness (e.g., more prominent inclusion of home oxygen requirements, enteral-feeding dependence, consultant recommendations, planned diagnostic follow-up, and clearer chronology) rather than fabricated diagnoses or fundamentally incorrect ICU courses.

### Cross-domain conflict detection

Because role-specialized agents generated independent views of the same patient, PAUSE-Agents could compare those views and surface contradictions for the receiving clinician rather than forcing the synthesis agent to silently choose one account. Across the 84 summative briefs, the resident agent surfaced 118 cross-domain conflict warnings (median 1 [IQR 0–2] per brief), of which 53 were safety-critical, 52 clinical, and 13 logistical (**Supplementary Table S8**); the safety-critical conflicts are cataloged thematically in **Table 2**. The deterministic safety layer raised an additional 421 safety flags (median 5 [IQR 4–6] per brief). Both surfaced once as clinician-facing WARNING banners on the editable draft for adjudication at transfer (**Supplementary Note S2**), rather than as repeated interruptive alerts; because these flags were uniformly severity-tagged, severity-based ranking to reduce reviewer burden is noted as future work (Discussion).

**Table 2.**
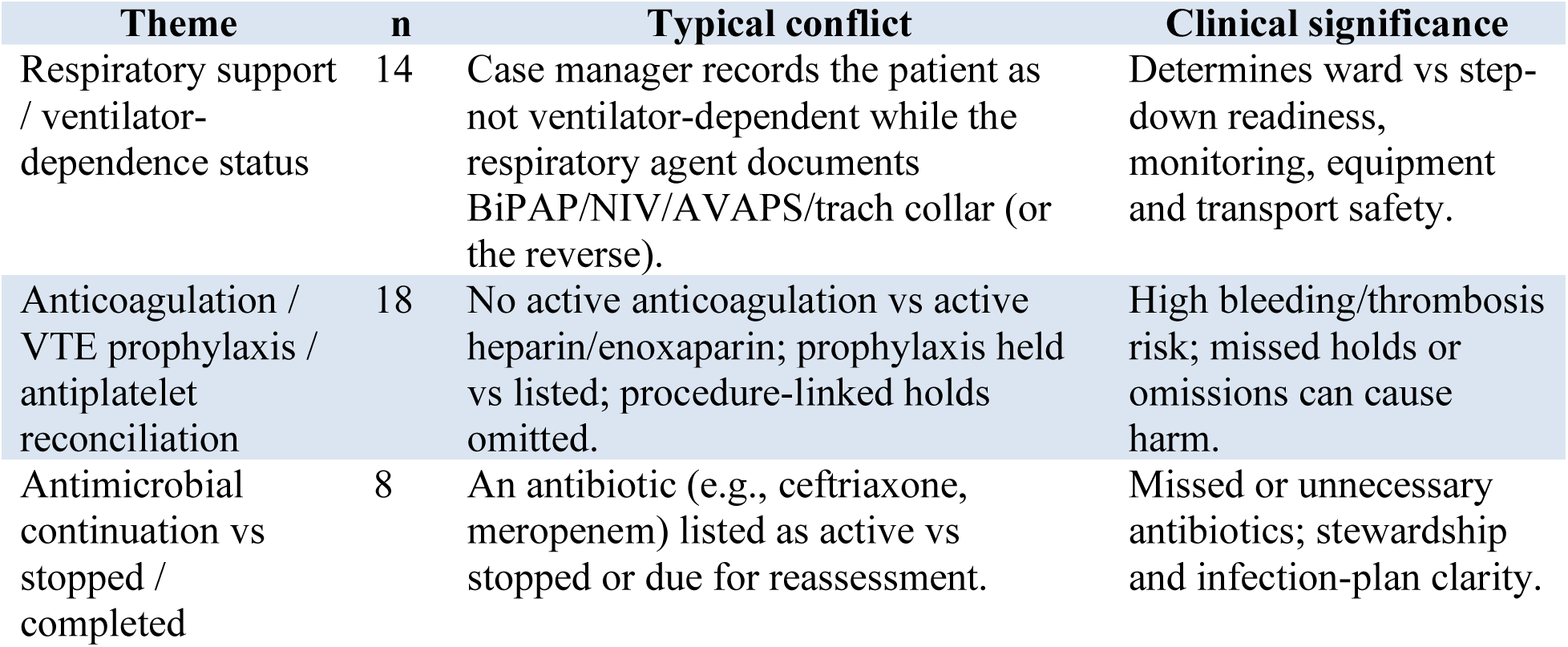
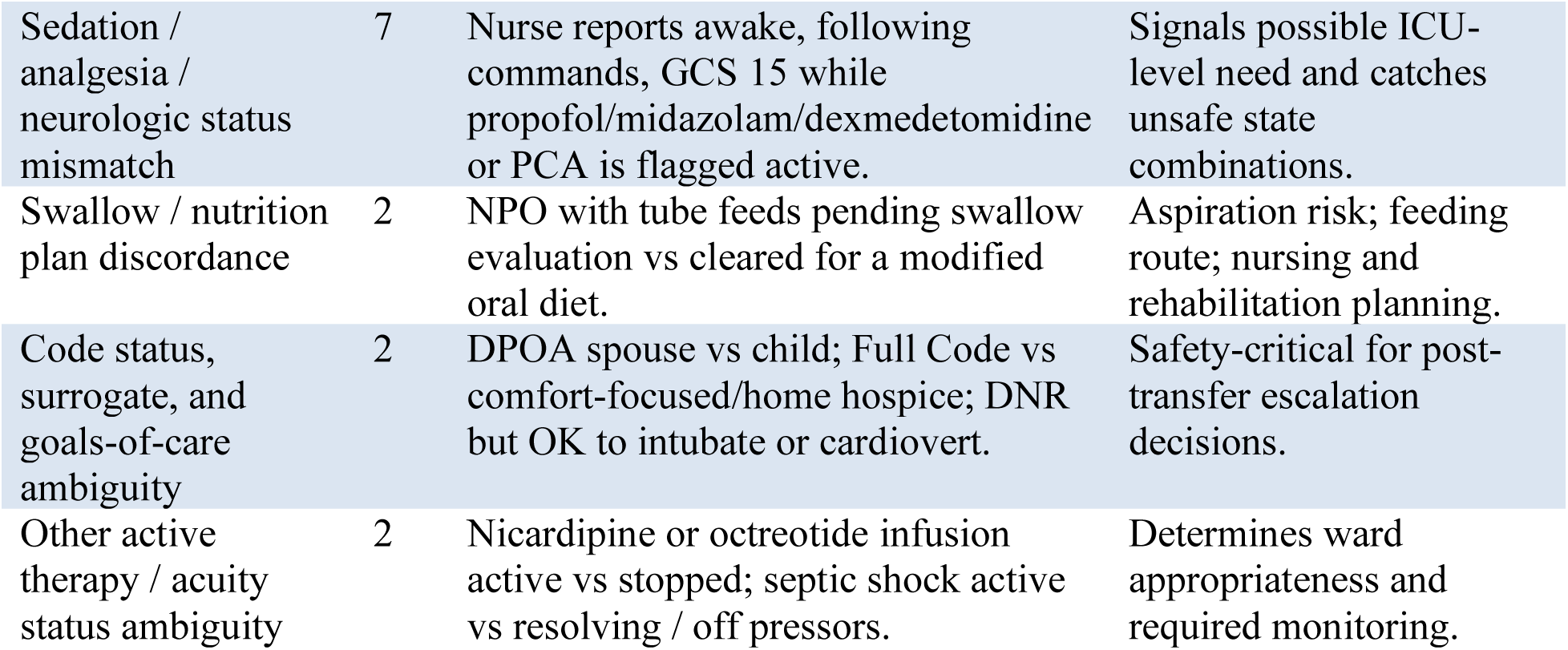
Thematic catalog of safety-critical cross-domain conflict warnings surfaced by PAUSE-Agents across 84-brief summative cohort. Themes were derived from the safety-critical cross-domain conflict warnings surfaced. Conflicts were paraphrased to remove identifiers. Severity follows the QA-banner scale described in Supplementary Note S2. The resident agent identifies the disagreeing domains, summarizes the conflict, assigns severity, and links the warning to the affected ICU PAUSE handoff section(s). PAUSE-Agents surface these conflicts for clinician review rather than resolving them automatically. BiPAP, bilevel positive airway pressure; NIV, noninvasive ventilation; AVAPS, average volume-assured pressure support; GCS, Glasgow Coma Scale; PCA, patient-controlled analgesia; NPO, nil per os; DPOA, durable power of attorney, DNR, do not resuscitate. Counts (n) reflect keyword-assigned themes over the 84-brief cohort; the full severity breakdown across all 118 conflicts is in Supplementary Table S8.

| Theme | n | Typical conflict | Clinical significance |
| --- | --- | --- | --- |
| Respiratory support / ventilator-dependence status | 14 | Case manager records the patient as not ventilator-dependent while the respiratory agent documents BiPAP/NIV/AVAPS/trach collar (or the reverse). | Determines ward vs step-down readiness, monitoring, equipment and transport safety. |
| Anticoagulation / VTE prophylaxis / antiplatelet reconciliation | 18 | No active anticoagulation vs active heparin/enoxaparin; prophylaxis held vs listed; procedure-linked holds omitted. | High bleeding/thrombosis risk; missed holds or omissions can cause harm. |
| Antimicrobial continuation vs stopped / completed | 8 | An antibiotic (e.g., ceftriaxone, meropenem) listed as active vs stopped or due for reassessment. | Missed or unnecessary antibiotics; stewardship and infection-plan clarity. |
| Sedation / analgesia / neurologic status mismatch | 7 | Nurse reports awake, following commands, GCS 15 while propofol/midazolam/dexmedetomidine or PCA is flagged active. | Signals possible ICU-level need and catches unsafe state combinations. |
| Swallow / nutrition plan discordance | 2 | NPO with tube feeds pending swallow evaluation vs cleared for a modified oral diet. | Aspiration risk; feeding route; nursing and rehabilitation planning. |
| Code status, surrogate, and goals-of-care ambiguity | 2 | DPOA spouse vs child; Full Code vs comfort-focused/home hospice; DNR but OK to intubate or cardiovert. | Safety-critical for post-transfer escalation decisions. |
| Other active therapy / acuity status ambiguity | 2 | Nicardipine or octreotide infusion active vs stopped; septic shock active vs resolving / off pressors. | Determines ward appropriateness and required monitoring. |

Safety-critical conflicts clustered around handoff elements where unresolved ambiguity could plausibly affect ward readiness or immediate management. Common categories included respiratory support and ventilator-dependence status, anticoagulation or venous thromboembolism (VTE) prophylaxis status, code status, goals of care, surrogate decision-making, antimicrobial continuation, sedation or analgesia state, and nutrition route. These warnings are best interpreted as prospective review targets: they identify internally discordant representations in the chart that could affect transfer safety, not necessarily errors in the generated brief. This is the central architectural rationale for the multi-agent design. While a single model can be prompted to identify contradictions, it does not generate independently scoped specialty views whose disagreements can be audited; PAUSE-Agents makes that comparison explicit and preserves unresolved disagreements as reviewable handoff artifacts.

Physicians were permitted to comment on these warnings during a structured review. Reviewers who did so generally affirmed that the flags identified real and clinically relevant inconsistencies, with occasional feedback that a flag over-weighted an otherwise stable finding. Because this response was optional and reviewer-initiated rather than a structured per-flag adjudication, we report these comments as preliminary support for the warning mechanism rather than as a positive predictive value. Formal adjudication of safety-critical conflict warnings is a planned next step.

### Pipeline architecture and computational performance

The PAUSE-Agents pipeline (**Figure 1a**) routed each patient’s data through a structured-extraction scribe, 6 role-specialized clinical agents (**Table 3**), a deterministic safety/quality layer, and two-stage resident-then-intensivist synthesis before deterministic merge-and-rendering of the final brief. In the primary GPT-5.4 configuration, the pipeline consumed a median of 418,334 input tokens and produced 25,127 output tokens per patient. The generated brief reduced the median source-to-output token ratio by approximately 17-fold, producing an editable ICU-PAUSE draft that physicians reviewed in a median of 10.5 minutes per case during evaluation. Token use reflected the deliberately redundant multi-agent design, in which role-specialized agents independently read routed slices before synthesis; detailed per-agent token economy is provided in **Supplementary Figure S1**. The quality-check agent consumed a median of zero tokens because first-line checks were deterministic and invoked the LLM only when a semantic cross-check was triggered.

**Table 3.** Agents: clinical responsibility, role-scoped input data, and owned ICU-PAUSE sections. “Active medication states” denotes the output of the deterministic medication-state classifier rather than raw administration rows. ICU-PAUSE sections: I, ICU admission/course; C, code status/goals/point of contact; U-unprescribing, unprescribing and high-risk medications; P, pending tests; A, active consultants; U-uncertainty, uncertainty measures; S, summary of problems and to-dos; E, exam at transfer. ADT, admission–discharge–transfer; SOFA, Sequential Organ Failure Assessment; H&P, history and physical; PT/OT, physical/occupational therapy.

| Agent | Tier | Clinical responsibility | Input data (structured + notes) | Section(s) |
| --- | --- | --- | --- | --- |
| Scribe | Extraction | Structured extraction of past medical history, allergies, home medications, code status, and admission-antibiotic courses; each field self-validated against the source notes | Routed clinical notes (H&P, progress, consult) | Pins → I, U-unprescribing, C |
| Nurse | Specialist | Vitals trends, hemodynamic and sedation state, lines/drains/skin findings | Vitals, patient assessments, demographics, ADT, SOFA, active medication states, notes | S, E |
| Respiratory Therapist | Specialist | Ventilator mode, weaning trials, secretion character, positioning | Respiratory support, vitals, labs, position, microbiology, notes | S, E |
| Pharmacist | Specialist | Medication reconciliation, renal/hepatic dose context, antibiotic-course tracking, unprescribing | Continuous/intermittent medications, labs, microbiology, notes + scribe antibiotic pins | U-unprescribing, S |
| Dietitian | Specialist | Nutrition status, albumin, feeding tolerance | Labs, vitals, patient assessments, notes (medications excluded — no nutrition-admin data in CLIF v2.1) | S |
| Case Manager | Specialist | Disposition, code status, psychosocial and discharge planning | Code status, ADT, demographics, notes | C, A, S |
| Rehab (PT/OT) | Specialist | Mobility, functional independence, fall risk, rehabilitation potential | Patient assessments, position, notes | A, S, E |
| Quality Check | Review | Deterministic safety/consistency gate, then optional LLM cross-check | All domain-agent snippets + full structured record | — (validation) |
| Resident | Review | Pre-synthesis review naming cross-domain conflicts, gaps, and priority to-dos | Specialist snippets and QA issues only (no raw record) | Pre-brief |
| Intensivist | Synthesis | Attending-level review of conflicts, gap-filling, and authoring of the full note | All snippets + full structured record + scribe pins + deterministic pending-tests block | I, S, E, C, A, P, U-unprescribing, U-uncertainty |

#### Evaluation efficiency

Physician review was the rate-limiting step of evaluation: reviewers spent a median of 10.5 minutes per case (IQR, 7.2-17.1; n = 100 reviews), totaling approximately 26 clinician-hours across the cohort. This evaluator burden was orders of magnitude greater than brief generation or automated scoring, motivating the LLM-as-a-Judge analysis below. Time-on-task reflects evaluation effort rather than brief quality.

### Automated PDSQI-9 judge as an exploratory quality monitor

#### Criterion validity

Across the 84 summative briefs, the o4-mini’s accuracy score correlated in the expected direction with physician-counted incorrect claims per brief (Spearman ρ = -0.29 [95% CI, - 0.49 to -0.08]; Figure 4a). The Thorough-omission axis was null, largely because 88% of briefs (74/84) had zero physician-flagged pertinent omissions, leaving little variance to track. Because the GPT-5.4 briefs were uniformly high quality, both physician error counts and judge scores were range-restricted, so these correlations should be interpreted as attenuated lower bounds.

**Figure 4.**
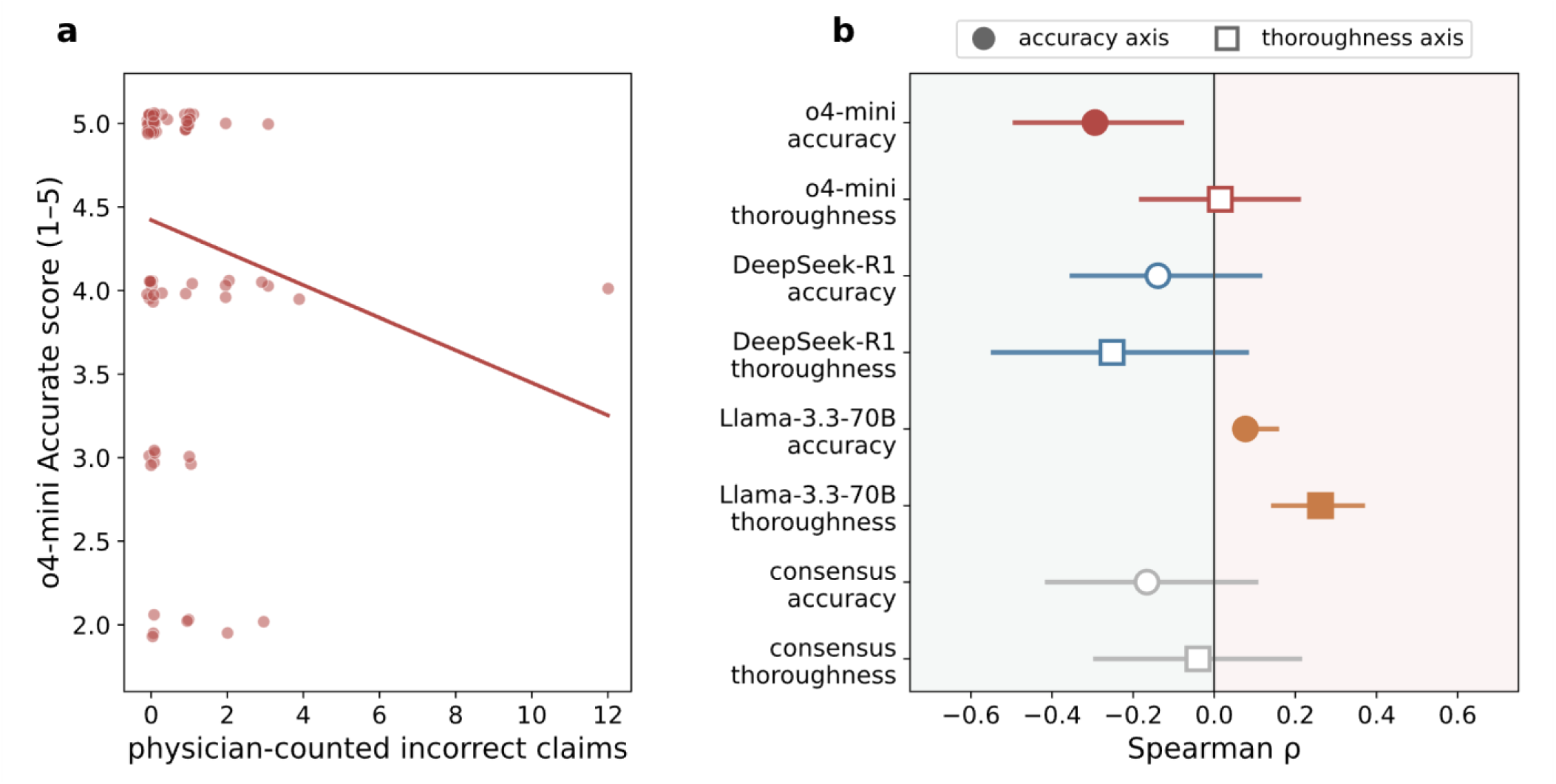
Criterion validity of the automated PDSQI-9 judge accuracy against physician review. Automated PDSQI-9 judges were evaluated on the 84 GPT-5.4 PAUSE-Agents briefs. (a) Association between physician-counted incorrect claims and the o4-mini PDSQI-9 Accuracy score. Each point represents one brief; the fitted line is shown only to visualize the overall trend. Briefs with more physician-identified incorrect claims tended to receive lower o4-mini Accuracy scores (Spearman ρ = −0.29; 95% CI, −0.49 to −0.08; n = 84 briefs). (b) Continuous criterion validity across candidate judges, measured as Spearman ρ between judge scores and physician objective error counts. Circles show the Accuracy axis, comparing judge Accuracy scores with physician-counted incorrect claims; squares show the Thoroughness axis, comparing judge Thoroughness scores with physician-counted pertinent omissions. Negative correlations indicate the expected direction, whereas positive correlations indicate wrong-signed associations. Points show correlations and horizontal bars show 95% bootstrap CIs; filled markers indicate CIs excluding zero. Only o4-mini Accuracy showed a correctly signed association with a CI excluding zero. Other judges and the equal-weight consensus were non-significant or wrong-signed. Because the GPT-5.4 briefs were uniformly high quality, these estimates reflect a range-restricted validation setting. AUROC and agreement analyses are shown in Supplementary Figure S2.

#### Discrimination and Agreement

The o4-mini accuracy score separated briefs with at least one physician-flagged incorrect claim from clean briefs with AUROC=0.58, indicating limited case-level discrimination despite being the strongest of the candidate judges. This performance is insufficient for safety clearance of individual briefs. We, therefore, interpret the judge as a screening tool for aggregate quality surveillance (including drift monitoring, model comparison, and prioritizing samples for physician audit) rather than as a replacement for clinician review. Agreement metrics alone were not sufficient: DeepSeek-R1 and Llama-3.3-70B achieved high AC2 values (0.97 and 0.96) but discriminated near chance on the accuracy axis (AUROC 0.51 and 0.49), whereas o4-mini had slightly lower AC2 (0.94) but the strongest discrimination. This indicates that the agreement on a compressed 4-5 score distribution can overstate criterion validity when brief quality is near ceiling (**Supplementary Figure S2**).

#### Efficiency

Automated PDSQI-9 scoring was inexpensive and parallelizable once briefs were generated, whereas physician review required a median of 10.5 minutes per case. We therefore view the judge as useful for aggregate quality surveillance, drift monitoring, and prioritizing cases for physician audit, but not as a substitute for clinician review of safety-critical outputs.

### Exploratory cross-model generalizability of the frozen pipeline

As an exploratory demonstration of scalability, we held the pipeline fixed and regenerated all 84 briefs with 3 open-weight generators (Qwen-3.6-27B, Gemma-4-31B, and MedGemma-27B) scoring each with the o4-mini judge (**Figure 5; Supplementary Table S6**).^29–31^ Because these comparisons used an automated judge with modest criterion validity rather than physician adjudication, they are relative, judge-screened signals rather than clinical error rates. Qwen-3.6 was closest to GPT-5.4 on several quality dimensions, whereas the medically fine-tuned MedGemma underperformed the general Gemma-4 model within this pipeline context. Detailed effect sizes, robustness controls, extraction-failure handling, and budget-parity checks are reported in **Supplementary Table S6.**

**Figure 5.**
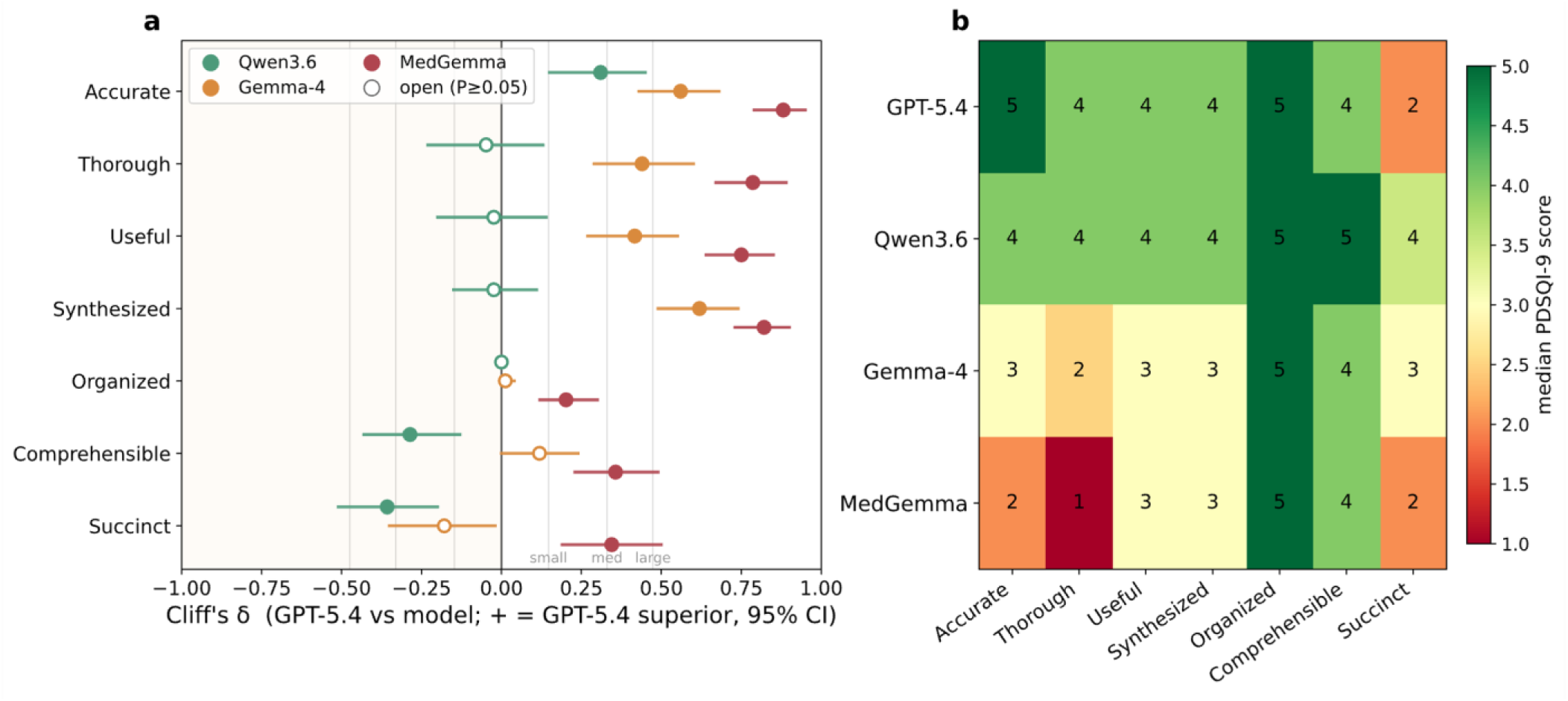
Cross-model generalizability of PAUSE-Agents (84 summative briefs; o4-mini PDSQI-9 judge). The PAUSE-Agents pipeline was held fixed and each base model regenerated all 84 briefs: GPT-5.4 (deployed reference), Qwen3.6-27B (Qwen3.6), Gemma-4-31B (Gemma-4), and the medically fine-tuned MedGemma-27B (MedGemma); full model identifiers in Supplementary Table S7. (a) Paired Cliff’s δ comparing GPT-5.4 with each open model across the seven PDSQI-9 content/readability dimensions (paired per hospitalization; 95% bootstrap CIs). Positive δ indicates GPT-5.4 superiority; open markers denote non-significant differences (P ≥ 0.05, Benjamini–Hochberg corrected). Vertical guides mark the negligible/small/medium/large effect-size thresholds (|δ| = 0.147 / 0.33 / 0.474). (b) Median PDSQI-9 score (1–5) by model and dimension. Because these comparisons use an automated judge with limited case-level discrimination, they are exploratory, judge-screened signals rather than physician-adjudicated error rates.

## Discussion

In this single-center development and evaluation study, PAUSE-Agents generated source-attributed ICU-to-ward handoff briefs that physicians judged accurate, complete, and useful as first drafts. Across 84 held-out MICU handoffs and 100 physician reviews, 98.8% of adjudicable claims were verified against the source record, pertinent omissions were uncommon, and briefs rated highly for organization, thoroughness, usefulness, and synthesis. At the brief level, 62% of briefs were entirely free of incorrect claims, while 38% contained at least one error requiring clinician review before use. This gap is the central rationale for our clinician-in-the-loop design. PAUSE-Agents produces a high-quality first draft, but a responsible clinician must still review, edit, and sign the final note before use.

The central contribution is architectural: PAUSE-Agents matches the multidisciplinary structure of critical care. ICU handoffs are not merely long-context summarization tasks; they require reconciling medication state, respiratory trajectory, code status, functional status, nutrition, disposition, pending diagnostics, and bedside exam. A role-specialized design makes these perspectives explicit before synthesis. When perspectives disagree, the disagreement becomes an output for clinician adjudication rather than an unobserved internal compromise by a single model. This distinction is central to the rationale for PAUSE-Agents and should be tested prospectively as a safety and workflow affordance, not only as a documentation-quality score.

The deterministic safety layer is equally important.^32^ High-stakes documentation should not rely on generative reasoning for facts that can be checked by rule. Medication activity, drug-drug interactions, laboratory thresholds, device dwell times, pending results, and numeric fidelity are better handled by auditable software tools that annotate or constrain LLM output. This deterministic-first design also makes errors easier to audit after deployment because safety flags, citation tags, and agent outputs can be traced back to their source rows and notes.

The marginal cost of operating the pipeline is likely small relative to the engineering, governance, and validation costs of deployment. Generating one brief consumed a median of approximately 418,000 input and 25,000 output tokens, dominated by repeated role-specialized reads of the chart; the corresponding per-brief API cost, with pricing date and rate assumptions, is reported in **Supplementary Note S6.** This cost is likely acceptable for a system intended to reduce handoff-preparation burden and surface discrepancies for review, although formal time-saved and cost-effectiveness analyses require prospective evaluation. The larger adoption barrier is mapping local EHR data into the pipeline input format and validating performance prospectively at each site.

The automated PDSQI-9 judge partially addresses the evaluation bottleneck. Physician review required substantial clinician time, whereas automated scoring was inexpensive and parallelizable. However, the judge should be interpreted as a monitoring screen rather than a replacement for human review. Its accuracy score was correlated in the expected direction with physician-counted errors, but discrimination was modest, and the validation corpus had restricted variance because the GPT-5.4 briefs were uniformly high quality. Future validation should include a physician-adjudicated sample spanning stronger and weaker generator outputs so that the judge is tested across a clinically meaningful quality range. To lower the barrier to such external validation, we release both the generation pipeline and the PHI-compliant clinician review application as open tools. Because the pipeline operates on the open CLIF common data model, other groups can map their own ICU data, regenerate briefs, and run the same 5-step human evaluation at their site rather than rebuilding the evaluation harness from scratch.

Several limitations warrant consideration. First, this was a single-center MICU study. Although PAUSE-Agents operates on CLIF, which has been adopted across multiple health systems,^26,33^ cross-institution portability has not yet been empirically demonstrated. Second, most summative briefs were reviewed by 1 physician, with overlap concentrated in targeted anchor cases for inter-rater reliability. Third, the 48-hour transfer anchor depended on identifying transfer notes by regular expression; chart review of 20 anchors showed 100% precision, but larger validation with a recall estimate remains future work.

Fourth, chronic oral-medication coverage was limited by source-data availability, contributing to some medication-related omissions. Fifth, the reviewer application displayed Philter-redacted notes, which produced some cannot-verify labels for durable power of attorney, point-of-contact, and other identity-dependent fields; this conservative review artifact would not apply to deployment in an unredacted HIPAA-compliant clinical environment. Finally, this initial evaluation did not include a direct comparator against clinician-authored handoffs or a single-pass, non-agentic LLM baseline, so we cannot isolate the incremental contribution of role specialization, deterministic checks, or cross-domain conflict surfacing.

Future work should move from retrospective first-draft quality to prospective clinical impact. The next evaluation should measure clinician editing burden, time saved, acceptance of source citations and warnings, alert fatigue from QA banners, adjudicated positive predictive value of cross-domain conflict flags, and downstream effects on transfer-note completeness and receiving-team situational awareness. Prospective comparisons against clinician-authored handoffs and single-pass LLM baselines will be needed to isolate the contribution of the multi-agent architecture. Because residual errors concentrated in ICU-course synthesis and medication reconciliation, future development should target temporal event ordering, medication-state reconciliation, and integration of outpatient medication lists, admission/discharge reconciliation, and pre-ICU MAR history.^34,35^ Because it already runs on a standardized intermediate format (CLIF), adding an ingestion layer based on HL7 Fast Healthcare Interoperability Resources (FHIR) –mapping FHIR resources into the retriever, or querying them directly through SMART on FHIR and bulk-FHIR APIs –is a natural next step toward EHR-native, multi-site deployment.^36,37^ PAUSE-Agents demonstrates that clinician-in-the-loop multi-agent LLM systems can generate high-quality, source-attributed ICU handoff briefs while preserving clinician oversight, and we release the complete pipeline and clinician evaluation platform to accelerate reproducible research and real-world implementation.

## Methods

### Study design and cohort

This was a single-center retrospective development and evaluation study of PAUSE-Agents, an LLM pipeline for ICU-to-ward transfer summarization, in the Northwestern Memorial Hospital medical intensive care unit (MICU). The corpus was partitioned into a development/human-in-the-loop (HITL) set of 25 ICU-to-ward transfer notes used for iterative refinement across 5 pilot review rounds and a held-out summative set of 84 notes used for final evaluation after the pipeline was locked (**Table 1**). The two sets were patient-disjoint, enforced at the patient level because one hospitalization may generate more than one transfer. The study was reviewed by the Northwestern University Institutional Review Board (IRB) and granted a waiver of informed consent given its retrospective design and use of already-collected clinical data (NU IRB #STU00224967). This study is reported in accordance with the TRIPOD-LLM reporting guideline for studies using large language models (**Supplementary Table S9**).^38^

EHR data were transformed into CLIF v2.1, an open common data model for longitudinal ICU data.^26^ We included MICU admissions during calendar year 2024 and anchored each brief to the patient’s first ICU-to-ward transfer. Because transfer notes are not recorded as a distinct note type at our institution, we identified the transfer anchor by regular-expression search over progress notes and used the matched note’s creation timestamp as the reference time. Source notes entered the bundle only if their creation timestamp preceded the reference time, preventing leakage from the transfer note itself. Each brief used data from the 48 hours before the reference time, with admission-stable notes such as the history and physical retained regardless of age to preserve the admission narrative. An ICU attending physician reviewed approximately 20 regex-identified anchors and confirmed that all represented genuine ICU-to-ward transfer notes; broader recall validation remains a task for future work. Cases were sampled with stratification by ICU length of stay (short <2 d; medium 2 to <5 d; long >=5 d) to span clinical complexity.

### Pipeline architecture

Briefs followed the clinician-developed 8-element ICU-PAUSE handoff template: ICU admission reason and brief course; code status, goals of care, and point of contact; areas of uncertainty; pending tests and results; active consultants; unprescribing and high-risk medications; summary of major problems and to-do items; and exam at transfer. We kept the clinical framework unchanged and added features required for an AI-generated first draft: fixed section rendering, inline source-citation tags, clinician-facing quality-assurance (QA) banners, and structured-plus-notes provenance (**Supplementary Note S1**).

PAUSE-Agents was implemented as an ordered multi-step workflow with deterministic retrieval, role-scoped LLM agents, deterministic safety tools, and two-stage synthesis. A deterministic retriever assembled each patient’s CLIF tables and routed clinical notes to the relevant agents under the 48-hour lookback window, selecting rows and notes by fixed rules rather than model judgment. Before prompting, each structured domain was condensed into a compact, trend-preserving text representation: vital signs were summarized as per-signal medians in fixed 8-hour buckets ordered from oldest to newest, with minimum, maximum, and count; laboratories and continuous infusions were represented by their most recent values or doses; and analogous deterministic reductions were applied to other domains. This aggregation preserved clinically relevant temporal trajectories while bounding token counts, mitigating lost-in-the-middle effects, and enabling the frozen pipeline to run across both the primary GPT-5.4 configuration and smaller open-weight comparators.^39^

After deterministic retrieval and condensation, each case was routed to a structured-extraction scribe and six role-specialized clinical agents: nurse, respiratory therapist, pharmacist, dietitian, case manager, and rehabilitation. Each agent received only its declared input slices and returned a structured snippet containing ICU-PAUSE section contributions, confidence, and warnings. Each role agent then performed a single self-critique pass over its own output to check for unsupported values, temporal misattribution of historical findings to active sections, and missing required sections, emitting a revised snippet before handoff to the safety layer.

The scribe extracted a small set of high-value structured fields from the notes before the role agents ran; we refer to these validated fields as “pins.” Pins included past medical history, allergies, code status, and admission-antibiotic courses. They were extracted once, validated against the source text, and propagated verbatim to downstream agents so the same anchor facts appeared consistently across sections. Failed pins were dropped rather than propagated, trading a small omission risk against reduced fabrication. Role-scoped inputs and ICU-PAUSE section ownership are summarized in **Table 3**, with implementation and routing details in **Supplementary Note S2**.

Downstream, a deterministic-first safety layer handled elements better verified by rule-based checks than by generative reasoning, including medication state, high-risk drug interactions, laboratory-value contradictions, device dwell time, section coverage, numeric fidelity, microbiologic grounding, and required propagation of scribe-extracted pins. The QA agent applied these validators before invoking an LLM semantic cross-check, which was used primarily to identify contradictions across specialist outputs. An information-restricted resident agent then reviewed the specialist snippets and QA issues to surface conflicts, gaps, and priority to-dos. Finally, an intensivist synthesis agent authored the eight ICU-PAUSE sections using the specialist outputs, full structured record, scribe pins, and deterministic pending-tests block. A deterministic merger de-duplicated content, filtered non-actionable problems, computed respiratory states directly from structured source data when device type and settings were unambiguous, and otherwise deferred to the respiratory agent’s narrative assessment. The final rendered draft included source-citation tags and clinician-facing warnings. Detailed validator logic and QA-banner conventions are provided in **Supplementary Note S3**.

### Secure computing environment

All protected health information (PHI) remained within the institution’s Health Insurance Portability and Accountability Act (HIPAA)-compliant secure computing environment. The proprietary generator and judge used the institution’s HIPAA-compliant Azure OpenAI deployment under a Business Associate Agreement (BAA), while open-weight models were served on-premises via vLLM on 2 NVIDIA H100 80 GB GPUs, so PHI never left local infrastructure. The clinician review application ran in the same HIPAA-compliant environment, and displayed redacted clinical notes passed through Philter so reviewers did not see raw identifiers.^40^ The automated judge scored the unredacted BAA-covered environment because no human viewed those inputs and redacting only the judge’s data would have broken comparability with the production pipeline. This reviewer-only redaction explains some redaction based cannot-verify labels and biases human-judge concordance toward the null rather than inflating it.

Context-window capacity was the primary feasibility constraint for candidate judges: Mixtral-8×22B (64K context) overflowed on 73/84 briefs and was excluded; 131K-context local judges had incomplete coverage for the longest briefs; only the 200K-context o4-mini judge covered the full cohort.^41^ Two cross-family comparators that did fit were retained only as robustness checks rather than reported judges. DeepSeek-R1-Distill-Qwen-32B returned all-zero scores on 16/84 briefs and Llama-3.3-70B was wrong-signed on the criterion-validity test; thus, a single o4-mini judge is reported. Additional deployment and review-application details are in **Supplementary Note S4;** the full generator and judge inventory is in **Supplementary Table S7**, with exact model identifiers, deployment versions, and coverage counts are in **Supplementary Note S5**.

### Clinician evaluation

Evaluation proceeded in 2 phases. During HITL development, the same 5 physicians reviewed 25 development briefs across 5 pilot rounds, and their feedback drove pipeline refinement. In the locked summative phase, 5 physician reviewers (2 hospitalist attendings, 2 internal medicine residents, and 1 pulmonary critical care attending) completed 100 structured reviews of 84 held-out briefs. Eighty briefs were reviewed by 1 clinician, and 4 anchor briefs were reviewed by all 5 clinicians for inter-rater reliability. This design prioritized broad evaluation of the held-out cohort while preserving a small set of fully overlapped anchor cases to estimate inter-rater reliability.^42,43^

Reviewers used a PHI-compliant web application that displayed the generated brief alongside the structured source data and routed notes (**Figure 2**). For each case, reviewers completed a 5-step rubric: sentence-level accuracy, marking each atomic claim as verified, incorrect, or cannot verify; domain-level completeness across 9 clinical data domains; PDSQI-9 documentation-quality ratings using the validated anchors; feedback on surfaced QA warnings; and optional free-text comments.28 For claim-and omission-level analyses, all 100 physician reviews were included. For brief-level PDSQI-9 analyses, the 5 ratings for each anchor brief were averaged into a single per-brief value, so each of the 84 held-out briefs contributed once. Physician review was labor-intensive, with a median review time of 10.5 minutes per case, which motivated the exploratory automated judge analysis described below.

### Automated judge and cross-model analyses

To evaluate whether automated scoring could support scalable quality monitoring, an o4-mini LLM judge scored each summative brief with the same PDSQI-9 rubric used by physicians, following the LLM-as-a-Judge approach of Croxford et al.^28^ The prompt included the validated PDSQI-9 definitions and anchors, PAUSE-Agents citation-tag and QA-banner conventions, and a strict JSON output contract. Because the o-series API does not expose a fixed temperature and scores varied run-to-run, each brief was scored over 5 iterations, and the per-attribute median was used. DeepSeek-R1-Distill-Qwen-32B and Llama-3.3-70B-Instruct were evaluated as cross-family comparators; o4-mini was retained as the primary judge because it had the strongest criterion validity in this cohort.^44,45^ Judge-prompt details, parsing, comparator models, and range-restriction diagnostics are provided in **Supplementary Note S5**.

Judge validity was assessed against physician-counted errors and omissions. The primary criterion-validity tests were Spearman correlations between a) the *Judge Accuracy* score and the number of physician-flagged incorrect claims, and b) the *Judge Thorough* score and the number of pertinent omissions; a valid judge was expected to assign lower scores as objective errors increased. Same-item AUROC was used only as a directional discrimination check, not as evidence of case-level safety clearance. In an exploratory cross-model analysis, the frozen PAUSE-Agents graph regenerated all 84 briefs with three open-weight generators (Qwen-3.6, Gemma-4, and MedGemma), and outputs were screened by the o4-mini judge. Because this comparison was judge-screened rather than physician-adjudicated, it is interpreted as a scalability signal rather than a clinical error-rate comparison.

### Statistical analysis

Cohort characteristics were compared with the Mann-Whitney U test for continuous variables and Fisher’s exact test for sex. Physician inter-rater reliability on anchor briefs used Gwet’s AC2 with exact-agreement percentages; the intraclass correlation coefficient (ICC) was non-estimable owing to near-uniform anchor quality and is reported in the Supplement. Reviewer identity effects were estimated by the gain in R2 from adding reviewer fixed effects. Judge correlations used 2,000 bootstrap resamples for 95% confidence intervals, with partial Spearman correlation adjusting for brief length as a sensitivity analysis. Cross-model comparisons used paired per-hospitalization differences with Wilcoxon signed-rank tests and Benjamini-Hochberg correction across PDSQI-9 dimensions; Cliff’s δ and Hodges-Lehmann median differences summarize effect sizes. Analyses used Python 3.12 (SciPy 1.17.1, pingouin 0.6.1, statsmodels 0.14.6).^46–48^

## Data availability

The datasets used in this study are not openly accessible due to privacy and security considerations. Electronic health record (EHR) data cannot be freely redistributed to researchers outside Institutional Review Board–sanctioned collaborations with the institution. Researchers seeking access to the data for legitimate scientific purposes may submit a reasonable request to the corresponding authors. Access will be considered on a case-by-case basis and, if approved, will require appropriate institutional approvals, including execution of a Data Use Agreement and compliance with all applicable ethical and regulatory requirements.

## Code availability

The code for the PAUSE-Agents pipeline, including the clinician review application used for human evaluation, is available at https://github.com/amagais/pause-agents. The pipeline runs on data in the open Common Longitudinal ICU Format (CLIF) v2.1. A site can map its own EHR data into CLIF using the specification and tooling at https://clif-icu.com and then download and run the released code locally. All model inference can run on-premises via vLLM — the open-weight specialist agents, and optionally an open-weight judge (our validated quality monitor was the BAA-covered o4-mini; see Discussion) — so that no protected health information need to leave the institution.

## Acknowledgements

S.A. is supported by the American Heart Association (AHA) Predoctoral Fellowship (25PRE1372516). C.A.G. is supported by the National Institutes of Health (NIH)/National Heart, Lung, and Blood Institute (NHLBI) K23HL169815. J.R. is supported by the NIH/National Institute on Drug Abuse (NIDA) R01DA051464 and the Robert Wood Johnson Foundation. P.L. is supported by the NIH K08CA270383, unrelated to this work.

## Author contributions

SA, JR, CAG, GF, DL, AK, NJ, and YL contributed to conceptualization. SA designed, implemented, and troubleshot the PAUSE-Agents pipeline, with methodological input from JR, YL, and CAG. DL provided technical support for Azure deployment and infrastructure troubleshooting, as well as clinical expertise. SA, WL, and CAG contributed to data curation and investigation. CM, CR, YL, BA, and CAG conducted physician evaluation. LS and PL provided clinical expertise on ICU-to-ward transfer workflows and the ICU-PAUSE handoff template. JR, YL, and CAG provided supervision and project administration. SA drafted the manuscript, and all authors reviewed and edited the manuscript.

