## Supplementary Materials for "PAUSE-Agents: A Clinician-in-the-Loop Multi-Agent AI Pipeline for ICU-to-Ward Handoff Briefs"

#### **Supplementary Note S1.** ICU-PAUSE template adaptation

The generated briefs followed the clinician-developed ICU-PAUSE structure without changing the clinical intent of the mnemonic. The eight rendered sections were: I, ICU admission reason and brief course; C, code status, goals of care, and point of contact; U, areas of uncertainty or diagnostic pause; P, pending tests and results; A, active consultants, including rehabilitation; U, unprescribing and pertinent high-risk medications; S, summary of major problems and to-do items; and E, exam at transfer. The AI-specific additions were limited to rendering and reviewability: fixed section headers, inline source-citation tags, clinician-facing warning banners, and paired structured-data plus note provenance.

Inline citation tags use a compact source-and-time convention, for example "(lab 5-27 06:00)" or "(progress_note 5-27 14:23)," so the signing clinician can trace factual claims back to the displayed source bundle. The template is rendered as a first draft for clinician editing, not as an autonomous note. Warning banners are shown above the brief when the pipeline identifies safety-critical flags, cross-domain conflicts, or data gaps that should be reviewed before finalizing the handoff.

#### **Supplementary Note S2.** Pipeline implementation and routing

PAUSE-Agents was implemented as a directed acyclic graph in LangGraph, with structured-data transformations in Polars and inter-node states represented as a Pydantic-validated typed object. Each agent used a versioned YAML prompt and returned a structured snippet containing proposed content, source of references, confidence, and warnings. The graph used deterministic retrieval and rendering around LLM calls so that source filtering, temporal anchoring, citation registry construction, and final assembly remained auditable.

The deterministic retriever assembled hospitalization-filtered CLIF tables and routed notes relative to the ICU-to-ward transfer reference time. Datetimes were normalized, source rows were restricted to the 48-hour lookback with a strict upper bound at the reference time, and future results were masked as pending rather than treated as known. Admission-stable notes, especially the history and physical, were retained regardless of age to preserve the admission narrative. Notes were routed by type to role-relevant agents; the H&P was made broadly available to clinical agents because it anchors the reason for ICU admission.

Structured CLIF domains were condensed before prompting. High-frequency vital signs were summarized into fixed 8-hour median buckets, with empty early buckets shown explicitly. Laboratories were de-duplicated within the window. Medications were converted into medication-state summaries rather than raw administration rows. Respiratory support, assessments, microbiology, procedures, code status, and ADT context were rendered into compact role-specific text. This role-scoped condensation kept individual prompts within context limits while preserving temporal features needed for clinical reasoning.

The scribe acted as an extraction agent rather than a prose generator. It emitted structured pins for high-value fields including past medical history, allergies, home medications, code status, and admission-antibiotic courses. Pins were self-validated against source-note substrings; pins that failed validation were dropped rather than passed downstream. Validated pins were injected into downstream inputs, especially the pharmacist and intensivist agents, to provide source-grounded anchors for medication and global synthesis.

Six specialist agents then produced independent role-scoped views: nurse, respiratory therapist, pharmacist, dietitian, case manager, and rehabilitation. An information-restricted resident agent reviewed only the specialist snippets and QA issues, not the raw record, and produced a pre-synthesis review naming cross-domain conflicts, critical gaps, and priority to-dos. The intensivist synthesis agent received the specialist outputs, full structured record, scribe pins, and deterministic pending-tests block, then authored all eight ICU-PAUSE sections. The deterministic merger de-duplicated content, removed generic non-actionable problems, computed vent-dependency status directly from respiratory data where possible, and assembled the final brief. Generation settings: domain agents ran at temperature 0.2 (the structured-extraction scribe and other extraction agents at 0.0), top-p 1, presence and frequency penalties 0, and a 4,096-token output cap per agent (raised for the intensivist synthesis); all cloud inference used the Azure OpenAI API (API-version 2024-12-01-preview). The o4-mini judge, an o-series reasoning model, does not expose temperature or top-p controls and ran at its default sampling with a raised completion-token budget.

#### **Supplementary Note S3.** Deterministic safety tools and QA banners

The safety layer followed a deterministic-first design: facts that can be checked by rule were computed in software and used to annotate, constrain, or flag LLM output. The drug-drug interaction checker combined a clinician-curated table of high-severity ICU-relevant pairs with optional openFDA drug-label querying. The curated table uses canonical medication names, order-invariant matching, severity tags, and auditable evidence sources. The openFDA query was network-gated and limited to moderate-severity alerts, whereas high-severity medication flags were derived only from the curated ICU medication table.

Medication-state classification labeled medications as active, recently stopped, or historical using drug-specific and renal-status-aware dosing intervals, alias canonicalization, and a trending-to-zero override for weaned infusions. Candidate drugs for interaction checking were restricted to active medications. Additional deterministic validators flagged contradictions between qualitative laboratory statements and numeric values, lines or devices exceeding dwell-time thresholds, numeric-fidelity mismatches outside tolerance, unsupported microbiology claims, missing section coverage, and failure to propagate required scribe-extracted pins such as admission-antibiotic courses.

QA banners were clinician-facing warnings, not automatic corrections. The resident agent surfaced unresolved issues by naming the disagreeing domains, describing the conflict, assigning severity, and linking the issue to affected ICU-PAUSE sections. Severity used a three-level scale: safety-critical for ambiguities that could plausibly affect ward readiness, immediate management, or patient safety, such as ventilator-dependence, anticoagulation, or code-status discrepancies; clinical for conflicts affecting clinical understanding or the care plan but unlikely to cause immediate harm if unresolved, such as non-urgent medication reconciliation or framing of a stable finding; and logistical for disposition, documentation, or process issues, such as mobility level or placement discrepancies. Banners were sorted with safety-critical issues first. Severity was recorded independently of issue category, so cross-domain conflicts could carry any severity. Internal audit-only outputs, including editorial self-critiques and process notes, were not shown to clinicians.

#### **Supplementary Note S4.** Secure computing environment and clinician review application

All model inference remained inside the institution's HIPAA-compliant environment. Proprietary models, including the GPT-5.4 generator and o4-mini judge, were accessed through the institution's HIPAA-compliant Azure OpenAI deployment under a Business Associate Agreement. Open-weight models were served on-premises through vLLM on institutional GPU infrastructure, so PHI did not leave local infrastructure for those runs. The clinician review application was deployed in the same secureAzure environment, restricted to named reviewers through Microsoft Entra ID single sign-on and per-reviewer credentials, with responses stored in institution-controlled Azure Blob Storage.

For reviewer display, clinical notes were passed through Philter so human reviewers did not see raw identifiers. This conservative de-identification step affected verification of identity-dependent claims such as DPOA, point-of-contact, names, and phone numbers. The automated judge scored the original unredacted BAA-covered bundles because no human viewed those inputs and because redacting only the judge inputs would have broken comparability with the production pipeline. Since redaction removed information from the human-review side only, this asymmetry should bias human-judge concordance toward the null rather than inflate it.

The review interface displayed the generated PAUSE-Agents brief alongside the structured source data and routed clinical notes. The source panel exposed the same categories of information available to the agents, including demographics, vitals, laboratories, infusions, intermittent medications, respiratory support, assessments, diagnoses, microbiology, procedures, and routed notes by type. Reviewers completed the five-step rubric within the application, and all responses were stored as structured JSON keyed by reviewer and case.

#### **Supplementary Note S5.** LLM-as-a-judge and cross-model evaluation details

The automated evaluator used the same PDSQI-9 rubric completed by physicians. The judge prompt included the validated PDSQI-9 definitions and anchors, the PAUSE-Agents citation-tag convention, the meaning of QA banners, and instructions not to score review aids as clinical assertions. The prompt required strict JSON output. For reasoning models, “thinking” blocks were stripped before parsing so that only the structured score object was analyzed. The o-series judge did not expose fixed temperature controls through the Azure API, so each brief was scored over five iterations and the per-attribute median was used; local judges were run at temperature 0.

o4-mini was selected as the primary judge because recent work found reasoning models to agree more closely with physicians for clinical-summary quality scoring, and because o4-mini had the strongest criterion validity in this cohort. Cross-family comparators included DeepSeek-R1-Distill-Qwen-32B and Llama-3.3-70B-Instruct served locally through vLLM. GPT-5.4 self-scoring was evaluated with a self-preference flag and excluded from headline ranking. Mixtral-8x22B was rejected for context infeasibility because many briefs exceeded its context window.

Because GPT-5.4 briefs were uniformly high quality, PDSQI-9 scores compressed near ceiling and rank-agreement statistics were not sufficient to establish validity. The primary judge-validity analysis therefore used continuous criterion validity: Spearman correlations between judge Accurate score and physician-counted incorrect claims, and between judge Thorough score and physician-counted pertinent omissions. A valid judge should assign lower scores as errors or omissions increase. Same-item AUROC provided a directional discrimination check, but the observed AUROC was treated as insufficient for case-level safety clearance. AC2 against a leave-one-out reviewer consensus ceiling and ICC were supporting measures only, and an AC2-saturation contrast was used to show that high agreement on clustered 4-5 scores does not necessarily imply discriminative validity.

For cross-model generalizability, the retriever, renderer, de-identification, and agent graph were frozen and all 84 summative briefs were regenerated with GPT-5.4, Qwen-3.6, Gemma-4, and MedGemma. DeepSeek-R1 served only as a judge. Qwen-3.6 was run with thinking disabled. Each generated brief was scored by the o4-mini judge over five iterations with a raised completion-token budget. Open-weight models were compared with GPT-5.4 using paired per-hospitalization differences, Wilcoxon signed-rank tests with Benjamini-Hochberg correction across dimensions, Cliff's delta with hospitalization-bootstrap confidence intervals, and Hodges-Lehmann median differences.

Robustness controls included requiring an effective judge-iteration count of at least three, budget-parity rejudging of iterations that exceeded the initial token budget after score-budget invariance checks, and exclusion of catastrophic extraction-agent failures. To characterize failure modes, the judge's free-text rationales on the lowest-scoring briefs were reviewed as illustrative signals rather than independent confirmation; fabrication-related observations were corroborated by manual physician review where reported. These cross-model results are therefore best interpreted as exploratory scalability signals, not as clinician-adjudicated error rates.

#### **Supplementary Note S6.** Deployment cost and token economy

Per-brief inference cost is reported here for readers who need exact figures. Generating one brief consumed a median of approximately 418,000 input and 25,000 output tokens (per-agent breakdown in the token-economy table). These tokens are spread across the pipeline’s routed per-agent requests rather than a single long-context call, so they are billed at the standard (under-272k-context) tier. At Azure OpenAI GPT-5.4 list pricing as of June 2026, Global pay-as-you-go is $2.50 per 1M input and $15.00 per 1M output tokens. This is approximately $1.42 per brief ($1.05 input plus $0.38 output), or about $1.56 per brief under Data Zone residency pricing ($2.75 and $16.50 per 1M). Because handoff briefs need not be generated in real time, the Azure Batch API (a 50% discount at $1.25 and $7.50 per 1M) roughly halves the marginal cost to about $0.71 per brief. Because most input tokens are patient-specific and unique to each case, further reductions would come mainly from tighter retrieval or on-premises open-weight inference; prompt caching offers little benefit here, as it applies only to the shared instruction prefix rather than the per-patient data that dominates the token count.

| PDSQI-9 attribute | n | Mean | SD | Median |
|---|---|---|---|---|
| Organized | 84 | 4.44 | 0.81 | 5.0 |
| Thorough | 84 | 4.36 | 0.88 | 5.0 |
| Useful | 84 | 4.34 | 0.78 | 5.0 |
| Comprehensible | 84 | 4.27 | 0.86 | 4.8 |
| Synthesized | 84 | 4.26 | 0.80 | 4.0 |
| Accurate | 84 | 4.18 | 0.93 | 4.1 |
| Cited | 84 | 3.92 | 1.03 | 4.0 |
| Succinct | 84 | 3.83 | 0.96 | 4.0 |
| **Total (mean of 8 attributes)** | **84** | **4.20** | **0.68** | — |
| Stigmatizing language (binary, % flagged) | 84 | 0% | — | — |

**Supplementary Table S1 | Physician PDSQI-9 attribute ratings of the deployed GPT-5.4 briefs (secondary outcome).** Distribution (n, mean, SD, median) of each PDSQI-9 attribute across the 84 summative briefs, rated by the five physicians on the instrument's validated 1–5 Likert scale (higher is better); each multiply-reviewed note is collapsed to the mean of its reviewers' scores. Rows are ordered from highest to lowest mean, followed by the per-note total (mean of the eight Likert attributes) and the binary Stigmatizing-language criterion (flagged in 0% of briefs).

| ICU-PAUSE section | Claims (n) | Verified | Cannot verify | Incorrect | Accuracy (%) |
|---|---|---|---|---|---|
| I | 545 | 512 | 10 | 23 | 95.7 |
| C | 584 | 548 | 34 | 2 | 99.6 |
| U-uncertainty | 354 | 348 | 1 | 5 | 98.6 |
| P | 390 | 387 | 2 | 1 | 99.7 |
| A | 904 | 895 | 4 | 5 | 99.4 |
| U-unprescribing | 761 | 732 | 6 | 23 | 97.0 |
| S | 1,220 | 1,206 | 4 | 10 | 99.2 |
| E | 1,160 | 1,126 | 33 | 1 | 99.9 |
| **Total** | **5,918** | **5,754** | **94** | **70** | **98.8** |

**Supplementary Table S2 | Sentence-level claim verification by ICU-PAUSE section (primary accuracy outcome).** Every clinical assertion in the 84 summative briefs was adjudicated by a physician as verified, incorrect, or cannot verify; counts are pooled across all reviewer evaluations (5,918 claims) and listed in canonical ICU-PAUSE order. Section accuracy is verified / (verified + incorrect), excluding cannot-verify claims from the denominator. Errors concentrate in the most synthesis-dependent sections (ICU Admission & Course, High-Risk Medications), while the structured-data-anchored Exam and Pending-tests sections approach ceiling. ICU-PAUSE sections: I, ICU admission/course; C, code status/goals/point of contact; U-unprescribing, unprescribing and high-risk medications; P, pending tests; A, active consultants; U-uncertainty, uncertainty measures; S, summary of problems and to-dos; E, exam at transfer.

| Data domain | Pertinent omissions (notes) | Completeness (%) |
|---|---|---|
| Intermittent medications | 6 | 92.9 |
| Recent labs | 4 | 95.2 |
| Continuous medications | 1 | 98.8 |
| Code status / goals of care | 1 | 98.8 |
| Respiratory / ventilator status | 0 | 100 |
| Microbiology / cultures | 0 | 100 |
| Vital-sign trends | 0 | 100 |
| Active consultants | 0 | 100 |
| Pending procedures / tests | 0 | 100 |

**Supplementary Table S3 | Pertinent omissions by clinical data domain (primary completeness outcome).** For each of the 9 data domains reviewers assessed, the number of summative briefs in which a clinically pertinent omission was flagged and the corresponding per-domain completeness (100 − the pertinent-omission rate across the cohort). Domains are ordered from lowest to highest completeness; the residual omissions concentrate in the intermittent-medication and laboratory domains, while the remaining seven domains are at or near 100%.

| Measure | Unit (n) | Gwet's AC2 | % exact |
|---|---|---|---|
| Claim verification (verified / cannot-verify / incorrect) | 207 claims | 0.98 | 95.8 |
| Claim correctness (correct vs incorrect) | 207 claims | 0.99 | 98.8 |
| Omission (complete vs something-missing) | 36 domain-cells | 0.97 | 96.7 |
| PDSQI-9 (secondary; mean over the 8 Likert attributes) | per attribute | 0.77 | — |

**Supplementary Table S4 | Inter-clinician agreement on the primary and secondary outcome measures.** Agreement among the 5 physicians on the 4 anchor briefs that all of them reviewed, reported as Gwet's AC2 alongside the percentage of exactly concordant judgments. The intraclass correlation was non-estimable because the uniformly high-quality anchor briefs leave almost no between-brief variance (range restriction), so AC2 and % exact agreement are reported instead. The objective claim- and omission-level measures (primary outcomes) are near-perfect, whereas the subjective PDSQI-9 ratings (secondary) are lower and leniency-sensitive.

| Theme | Summary | Representative clinician comment |
|---|---|---|
| **Strengths** | Reviewers frequently praised the summaries' thoroughness, organization, transfer planning, and synthesis of complex ICU courses. | "Probably the most thorough, organized, well-reasoned ICU summary that I've reviewed yet in this final round." |
| **Clinical completeness** | Reviewers occasionally identified additional contextual information that could be surfaced more prominently. | "Home O2 not listed in ICU summary, very relevant info for accepting hospitalist." |
| **Information prioritization** | Some reviewers suggested reordering or condensing information to better reflect clinical importance. | "Laryngectomy coming before Sepsis and Nutrition is not ordered by relevance." |
| **Diagnostic synthesis** | A small number of comments focused on how consultant conclusions or resolved diagnoses were incorporated into the narrative. | "Mentioning concern for a major artery occlusion without mentioning the follow-up eval by vascular surgery seems like a missed opportunity for synthesis." |
| **Timeline interpretation** | Some summaries contained ambiguities regarding the timing or location of major clinical events. | "The cardiac arrest happened before ICU." |
| **Medication reconciliation** | Reviewers occasionally identified discrepancies regarding active versus discontinued medications. | "Meropenem is an active medication per multiple notes, but is described as a prior med in the text summary." |
| **Communication of uncertainty** | Reviewers valued explicit discussion of unresolved questions and highlighted opportunities for clarification when uncertainty remained. | "I would want to clarify if I was taking over this patient to confirm they weren't on HD and didn't have a temp HD line." |
| **Anticipatory guidance and safety planning** | Reviewers highlighted several examples where the generated summaries proactively identified potential sources of harm or management confusion. | "Really good synthesis and pre-emptive discussion of a potential source of harm." |

**Supplementary Table S5 | Themes from the optional clinician free-text feedback.** Qualitative themes distilled from the free-text comments completed by clinicians during the evaluation of PAUSE-Agents generated handoff briefs, each paired with representative clinician feedback in quotations.

| Comparison | Dimension | Cliff's δ (95% CI) | Hodges–Lehmann Δ | P (BH) | δ (excl. extraction failures) |
|---|---|---|---|---|---|
| GPT-5.4 vs Qwen-3.6 | Accurate | +0.31 [+0.15, +0.45] | +0.50 | $1.2 \times 10^{-4}$ | +0.30 |
| GPT-5.4 vs Qwen-3.6 | Thorough | −0.05 [−0.23, +0.13] | 0.00 | 0.970 | −0.09 |
| GPT-5.4 vs Qwen-3.6 | Useful | −0.02 [−0.20, +0.14] | 0.00 | 0.839 | −0.05 |
| GPT-5.4 vs Qwen-3.6 | Synthesized | −0.02 [−0.15, +0.11] | 0.00 | 0.970 | −0.04 |
| GPT-5.4 vs Qwen-3.6 | Organized | +0.00 [+0.00, +0.00] | 0.00 | n.s. | +0.00 |
| GPT-5.4 vs Qwen-3.6 | Comprehensible | −0.29 [−0.43, −0.13] | −0.50 | $9.4 \times 10^{-4}$ | −0.30 |
| GPT-5.4 vs Qwen-3.6 | Succinct | −0.36 [−0.51, −0.20] | −0.50 | $1.2 \times 10^{-4}$ | −0.38 |
| GPT-5.4 vs Gemma-4 | Accurate | +0.56 [+0.43, +0.68] | +1.00 | $8.7 \times 10^{-9}$ | — |
| GPT-5.4 vs Gemma-4 | Thorough | +0.44 [+0.29, +0.60] | +1.00 | $1.9 \times 10^{-6}$ | — |
| GPT-5.4 vs Gemma-4 | Useful | +0.42 [+0.27, +0.55] | +0.50 | $1.2 \times 10^{-5}$ | — |
| GPT-5.4 vs Gemma-4 | Synthesized | +0.62 [+0.49, +0.74] | +0.50 | $3.8 \times 10^{-10}$ | — |
| GPT-5.4 vs Gemma-4 | Organized | +0.01 [+0.00, +0.04] | 0.00 | 0.363 | — |
| GPT-5.4 vs Gemma-4 | Comprehensible | +0.12 [+0.00, +0.24] | 0.00 | 0.078 | — |
| GPT-5.4 vs Gemma-4 | Succinct | −0.18 [−0.35, −0.02] | 0.00 | 0.078 | — |
| GPT-5.4 vs MedGemma | Accurate | +0.88 [+0.79, +0.95] | +2.50 | $8.8 \times 10^{-14}$ | — |
| GPT-5.4 vs MedGemma | Thorough | +0.79 [+0.67, +0.89] | +2.00 | $6.6 \times 10^{-13}$ | — |
| GPT-5.4 vs MedGemma | Useful | +0.75 [+0.64, +0.85] | +1.50 | $4.3 \times 10^{-12}$ | — |
| GPT-5.4 vs MedGemma | Synthesized | +0.82 [+0.73, +0.90] | +1.00 | $6.7 \times 10^{-14}$ | — |
| GPT-5.4 vs MedGemma | Organized | +0.20 [+0.12, +0.30] | 0.00 | $1.8 \times 10^{-4}$ | — |
| GPT-5.4 vs MedGemma | Comprehensible | +0.36 [+0.23, +0.49] | +0.50 | $8.9 \times 10^{-6}$ | — |
| GPT-5.4 vs MedGemma | Succinct | +0.34 [+0.19, +0.50] | +0.50 | $2.0 \times 10^{-4}$ | — |
| Gemma-4 vs MedGemma | Accurate | +0.70 [+0.57, +0.82] | +1.50 | $<10^{-3}$ | — |
| Gemma-4 vs MedGemma | Thorough | +0.57 [+0.44, +0.70] | +1.00 | $<10^{-3}$ | — |
| Gemma-4 vs MedGemma | Useful | +0.64 [+0.52, +0.75] | +1.00 | $<10^{-3}$ | — |
| Gemma-4 vs MedGemma | Synthesized | +0.29 [+0.15, +0.40] | +0.25 | $<10^{-3}$ | — |
| Gemma-4 vs MedGemma | Organized | +0.19 [+0.11, +0.29] | 0.00 | $<10^{-3}$ | — |
| Gemma-4 vs MedGemma | Comprehensible | +0.29 [+0.15, +0.42] | +0.50 | $<10^{-3}$ | — |
| Gemma-4 vs MedGemma | Succinct | +0.46 [+0.30, +0.62] | +1.00 | $<10^{-3}$ | — |

**Supplementary Table S6 | Cross-model generalizability: per-dimension paired effect sizes (o4-mini PDSQI-9 judge, 84 paired briefs).** Each open-weight generator (top three groups) is compared to the GPT-5.4 reference (positive Cliff's δ = GPT-5.4 superior); the final group is the direct Gemma-4-vs-MedGemma contrast establishing that medical fine-tuning degraded its own base model (positive δ = Gemma-4 superior). Hodges–Lehmann Δ is the median paired difference; P-values are Benjamini–Hochberg–corrected across the seven Likert dimensions; δ is also reported with catastrophic-extraction-failure cases excluded where computed.

| Model | Size | HF ID / provider | Context window | Role in study |
|---|---|---|---|---|
| **GPT-5.4** | undisclosed (cloud) | Azure OpenAI | ~1 M | Multi-agent generator |
| Gemma-4-31B | 31 B | google/Gemma-4-31b-it | 256 K native (served 131 K) | Multi-agent generator |
| MedGemma-27B | 27 B (medical-specialized) | google/MedGemma-27b | served 131 K | Multi-agent generator |
| Qwen3.6-27B | 27 B | Qwen/Qwen3.6-27B | 256 K native (served 131 K) | Multi-agent generator |
| **o4-mini** | undisclosed (cloud) | Azure OpenAI | 200 K | Sole reported judge |
| DeepSeek-R1-Distill-Qwen-32B | 32 B | deepseek-ai/DeepSeek-R1-Distill-Qwen-32B | 131 K | Comparator, dropped |
| Llama-3.3-70B-Instruct | 70 B | meta-llama/Llama-3.3-70B-Instruct | 128 K | Comparator, dropped |
| Mixtral-8×22B | 8×22 B MoE | mistralai/Mixtral-8x22B-Instruct-v0.1 | 64 K | Rejected — context-too small |

**Supplementary Table S7 | Model inventory: generators and judges.** Every language model used in the study, grouped by role, with its parameter size, Hugging Face identifier or cloud provider, context-window limit, access mode (metered cloud API vs. localy served open weights), cost structure, and specific role. Generators: GPT-5.4 (production generator and human-evaluated anchor) plus the open-weight Gemma-4-31B, MedGemma-27B, and Qwen3.6-27B used in the cross-model generalizability analysis. Judges: o4-mini (sole reported judge), with DeepSeek-R1-Distill-Qwen-32B and Llama-3.3-70B-Instruct as cross-family comparators (dropped for wrong-signed or saturated validity) and Mixtral-8×22B rejected for insufficient context window.

| Conflict theme | Safety-critical | Clinical | Logistical | Total |
|---|---|---|---|---|
| Respiratory support / ventilator-dependence | 14 | 16 | 7 | 37 |
| Anticoagulation / VTE prophylaxis / antiplatelet | 18 | 2 | 0 | 20 |
| Antimicrobial continuation vs stopped | 8 | 11 | 1 | 20 |
| Sedation / analgesia / neurologic status | 7 | 5 | 1 | 13 |
| Swallow / nutrition plan | 2 | 12 | 2 | 16 |
| Code status / surrogate / goals-of-care | 2 | 0 | 0 | 2 |
| Other active therapy / acuity status | 2 | 4 | 0 | 6 |
| Disposition / mobility / rehabilitation | 0 | 1 | 2 | 3 |
| Other / miscellaneous | 0 | 1 | 0 | 1 |
| **Total** | **53** | **52** | **13** | **118** |

**Supplementary Table S8. | Severity and thematic breakdown of the 118 cross-domain conflict warnings surfaced across the 84-brief summative cohort.** Each conflict the resident agent raised is cross-classified by clinical theme (rows) and severity (columns: safety-critical, clinical, logistical, following the QA-banner scale in Supplementary Note S2). Themes were assigned by keyword matching on the conflict text and reviewed for consistency; one warning did not map to a defined theme (Other / miscellaneous). Of the 118 warnings, 53 were safety-critical, 52 clinical, and 13 logistical, with anticoagulation/VTE-prophylaxis and respiratory/ventilator-dependence conflicts dominating the safety-critical column — which corresponds to the conflicts described qualitatively in Table 2 of the main text. This cross-domain signal cannot be produced by a single-pass summarizer by construction (see Results).

| Item | Checklist item | Research design | LLM task | Page |
|---|---|---|---|---|
| **1** | Identify the study as developing, fine-tuning, and/or evaluating the performance of an LLM, specifying the task, the target population, and the outcome to be predicted. | All | All | 1 |
| **2** | See TRIPOD-LLM for Abstracts. | All | All | 2 |
| **3a** | Explain the healthcare context / use case (e.g., administrative, diagnostic, therapeutic, clinical workflow) and rationale for developing or evaluating the LLM, including references to existing approaches and models. | All | All | 3 |
| **3b** | Describe the target population and the intended use of the LLM in the context of the care pathway, including its intended users in current gold standard practices (e.g., healthcare professionals, patients, public, or administrators). | E, H | All | 3 |
| **4** | Specify the study objectives, including whether the study describes the initial development, fine-tuning, or validation of an LLM (or multiple stages). | All | All | 3 |
| **5a** | Describe the sources of data separately for the training, tuning, and/or evaluation datasets and the rationale for using these data (e.g., web corpora, clinical research/trial data, EHR data, or unknown). | All | All | 18-19 |
| **5b** | Describe the relevant data points and provide a quantitative and qualitative description of their distribution and other relevant descriptors of the dataset (e.g., source, languages, countries of origin). | All | All | 6 |
| **5c** | Specifically state the date of the oldest and newest item of text used in the development process (training, fine-tuning, reward modeling) and in the evaluation datasets. | All | All | 18 |
| **5d** | Describe any data pre-processing and quality checking, including whether this was similar across text corpora, institutions, and relevant socio-demographic groups. | All | All | 6, 18 |
| **5e** | Describe how missing and imbalanced data were handled and provide reasons for omitting any data. | All | All | 6, 17-18 |
| **6a** | Report the LLM name, version, and last date of training. | All | All | Suppl Table S7 |
| **6b** | Report details of LLM development process, such as LLM architecture, training, fine-tuning procedures, and alignment strategy (e.g., reinforcement learning, direct preference optimization, etc.) and alignment goals (e.g., helpfulness, honesty, harmlessness, etc.). | M, D | All | N/A |
| **6c** | Report details of how text was generated using the LLM, including any prompt engineering (including consistency of outputs), and inference settings (e.g., seed, temperature, max token length, penalties), as relevant. | M, D, E | All | Suppl Note S2 |
| **6d** | Specify the initial and post-processed output of the LLM (e.g., probabilities, classification, unstructured text). | All | All | 12-13, 18-19 |
| **6e** | Provide details and rationale for any classification and, if applicable, how the probabilities were determined and thresholds identified. | All | C, OF | Suppl Note S3 |
| **7a** | Include metrics that capture the quality of generative outputs, such as consistency, relevance, and accuracy, compared to gold standards. | All | QA, IR, DG, SS, MT | 20-21 |
| **7b** | Report the outcome metrics' relevance to downstream task at deployment time and, where applicable, correlation of metric to human evaluation of the text for the intended use. | E, H | All | 20 |
| **7c** | Clearly define the outcome, how the LLM predictions were calculated (e.g., formula, code, object, API), the date of inference for closed-source LLMs, and evaluation metrics. | E, H | All | 18-21 |
| **7d** | If outcome assessment requires subjective interpretation, describe the qualifications of the assessors, any instructions provided, relevant information on demographics of the assessors, and inter-assessor agreement. | All | All | 18-21 |
| **7e** | Specify how performance was compared to other LLMs, humans, and other benchmarks or standards. | All | All | 15-17 |
| **8a** | If annotation was done, report how text was labeled, including providing specific annotation guidelines with examples. | All | All | 20 |

| | | | | |
|---|---|---|---|---|
| **8b** | If annotation was done, report how many annotators labeled the dataset(s), including the proportion of data in each dataset that were annotated by more than 1 annotator, and the inter-annotator agreement. | All | All | 14-15, 18 |
| **8c** | If annotation was done, provide information on the background and experience of the annotators or characteristics of any models involved in labelling. | All | All | 20 |
| **9a** | If research involved prompting LLMs, provide details on the processes used during prompt design, curation, and selection. | All | All | 18-21 |
| **9b** | If research involved prompting LLMs, report what data were used to develop the prompts. | All | All | 18 |
| **10** | Describe any preprocessing of the data before summarization. | All | SS | 18-19 |
| **11** | If instruction tuning/alignment strategies were used, what were the instructions, data, and interface used for evaluation, and what were the characteristics of the populations doing evaluation? | M, D | All | N/A |
| **12** | Report compute, or proxies thereof (e.g., time on what and how many machines, cost on what and how many machines, inference time, floating-point operations per second (FLOPs)), required to carry out methods. | M, D, E | All | Suppl Fig 1 |
| **13** | Name the institutional research board or ethics committee that approved the study and describe the participant-informed consent or the ethics committee waiver of informed consent. | All | All | 18 |
| **14a** | Give the source of funding and the role of the funders for the present study. | All | All | 22 |
| **14b** | Declare any conflicts of interest and financial disclosures for all authors. | All | All | 22 |
| **14c** | Indicate where the study protocol can be accessed or state that a protocol was not prepared. | H | All | N/A |
| **14d** | Provide registration information for the study, including register name and registration number, or state that the study was not registered. | H | All | N/A |
| **14e** | Provide details of the availability of the study data. | All | All | 22 |
| **14f** | Provide details of the availability of the code to reproduce the study results. | All | All | 22 |
| **15** | Provide details of any patient and public involvement during the design, conduct, reporting, interpretation, or dissemination of the study or state no involvement. | H | All | N/A |
| **16a** | When using patient/EHR data, describe the flow of text/EHR/patient data through the study, including the number of documents/questions/participants with and without the outcome/label and follow-up time as applicable. | E, H | All | 6-7, 18 |
| **16b** | When using patient/EHR data, report the characteristics overall and, for each data source or setting, and for development/evaluation splits, including the key dates, key characteristics, and sample size. | E, H | All | 6-7,18 |
| **16c** | For LLM evaluation that include clinical outcomes, show a comparison of the distribution of important clinical variables that may be associated with the outcome between development and evaluation data, if available. | E, H | All | 6-7 |
| **16d** | When using patient/EHR data, specify the number of participants and outcome events in each analysis (e.g., for LLM development, hyperparameter tuning, LLM evaluation). | E, H | All | 6-7 |
| **17** | Report LLM performance according to pre-specified metrics (see item 7a) and/or human evaluation (see item 7d). | All | All | 6-16 |
| **18** | If applicable, report the results from any LLM updating, including the updated LLM and subsequent performance. | All | All | N/A |
| **19a** | Give an overall interpretation of the main results, including issues of fairness in the context of the objectives and previous studies. | All | All | 16-18 |
| **19b** | Discuss any limitations of the study and their effects on any biases, statistical uncertainty, and generalizability. | All | All | 17-18 |
| **19c** | Describe any known challenges in using data for the specified task and domain context with reference to representation, missingness, harmonization, and bias. | E, H | All | 17-18 |
| **19d** | Define the intended use for the implementation under evaluation, including the intended input, end-user, level of autonomy/human oversight. | E, H | All | 3-4,16-18 |
| **19e** | If applicable, describe how poor quality or unavailable input data should be assessed and handled when implementing the LLM, i.e., what is the usability of the LLM in the context of current clinical care. | E, H | All | 17-18 |

| | | | | |
|---|---|---|---|---|
| **19f** | If applicable, specify whether users will be required to interact in the handling of the input data or use of the LLM, and what level of expertise is required of users. | E, H | All | 17-18 |
| **19g** | Discuss any next steps for future research, with a specific view to applicability and generalizability of the LLM. | All | All | 18 |

**Supplementary Table S9 | TRIPOD-LLM reporting checklist.** Completed the checklist from Galifant et al. (2025) to ensure transparency and reproducibility.

### Supplementary Figures

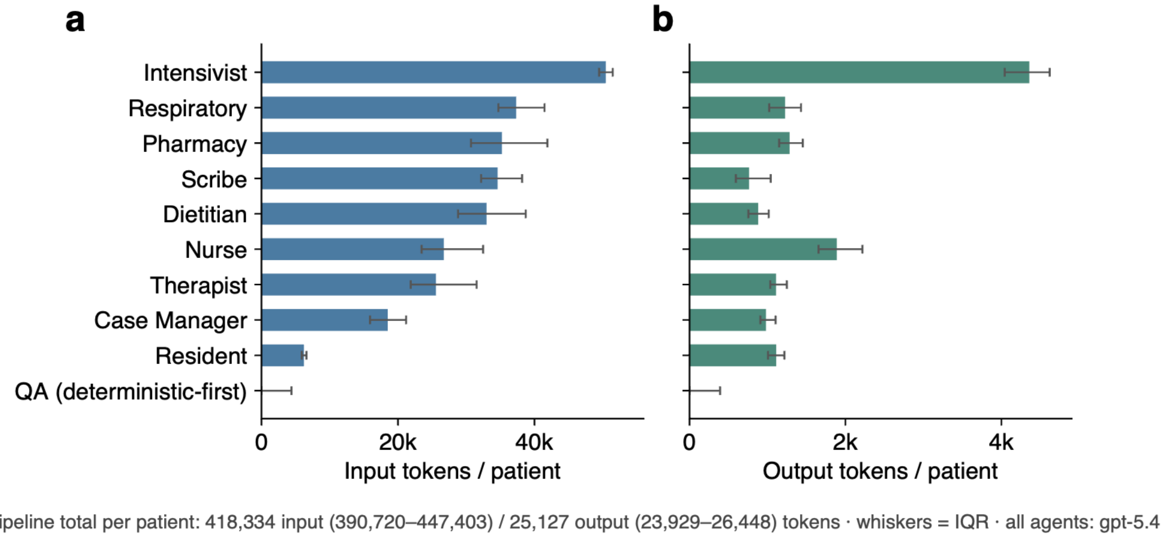


**Supplementary Figure S1 | Per-patient token economy by pipeline agent.** Median per-patient input (a) and output (b) tokens for each agent (whiskers = interquartile range); all agents run on GPT-5.4. The Intensivist synthesis agent has the largest input footprint because it reads every specialist snippet plus the full structured record, whereas the deterministic-first Quality-Check stage consumes near-zero generative tokens. Pipeline totals per patient: 418,334 input (IQR 390,720–447,403) and 25,127 output (IQR 23,929–26,448) tokens.

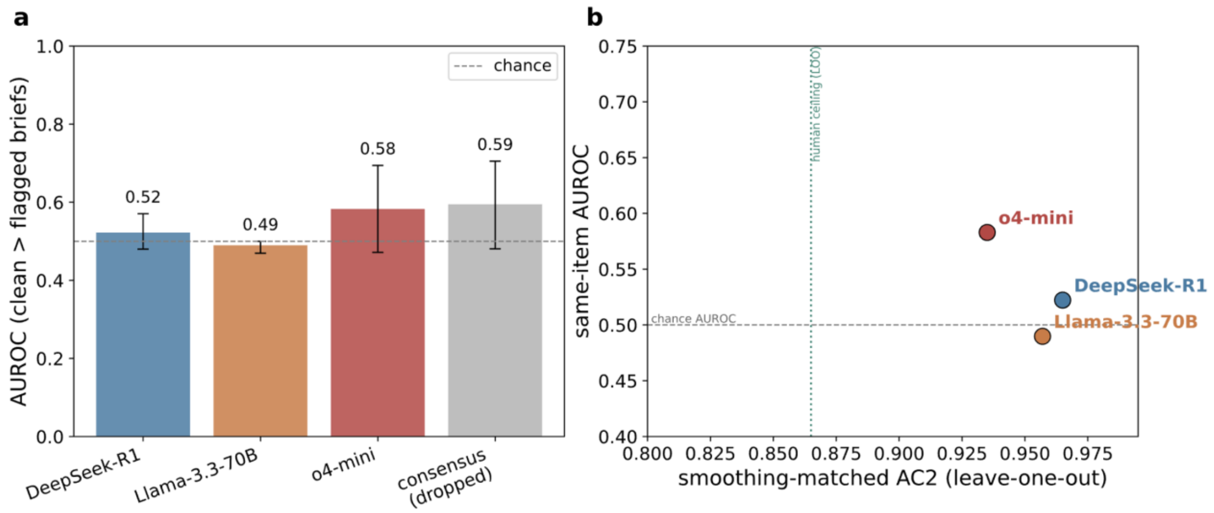


**Supplementary Figure S2.** Judge validation: discrimination versus agreement saturation. (a) Same-item accuracy discrimination — area under the ROC curve for separating clean from flagged briefs for each candidate LLM judge (dashed line = chance; the consensus blend is shown as dropped, difference not significant). (b) AC2 saturates: the highest inter-rater agreement (Gwet's AC2, x-axis; n = 4 anchor briefs, leave-one-out) does not coincide with the best same-item discrimination (AUROC, y-axis). o4-mini was retained as the deployed judge.
